# Genomic characterization of chikungunya virus during the large 2025 outbreak in Sri Lanka reveals emergence of a distinct Indian Ocean lineage strain

**DOI:** 10.1101/2025.05.23.25328206

**Authors:** Tibutius Thanesh Pramanayagam Jayadas, Malithi de Silva, Yunshi Liao, Feng Zhu, Esther Jinjin Zhang, Laksiri Gomes, Heshan Kuruppu, Ruwani Withanachchi, Radanee Rathnapriya, Farha Bary, Sahan Madusanka, Ruklanthi de Alwis, Tommy Tsan-Yuk Lam, Chandima Jeewandara, Gathsaurie Neelika Malavige

## Abstract

Chikungunya virus (CHIKV) has reemerged globally, with large outbreaks reported in multiple regions in 2025. Sri Lanka experienced a major outbreak after nearly two decades of minimal transmission. We aimed to characterize the genomic features of CHIKV responsible for this outbreak and compare them with historical and global strains. Acute febrile patients presenting between April and September 2025 were screened for CHIKV using quantitative PCR. Whole genome sequencing was performed using Oxford Nanopore technology. Phylogenetic, mutational, structural, and phylodynamic analyses were conducted using global reference datasets and integrated viral genomic, temporal, and geographical data.

All Sri Lankan sequences from 2025 clustered within the Indian Ocean lineage (IOL) of the East Central South African genotype, forming a distinct monophyletic clade. In contrast, outbreaks in Brazil, China, Réunion, and Mayotte were associated with genetically distinct lineages. The Sri Lankan and other South Asian strains were characterized by unique mutations in both structural and non-structural proteins. Key mutations in the E1 and E2 proteins were located at sites associated with mosquito vector competence, within the MXRA8 receptor binding interface, and in known neutralizing antibody binding regions. Compared with the 2006 to 2008 IOL epidemic strains, the 2025 viruses showed a markedly different molecular profile.

The 2025 chikungunya outbreak in Sri Lanka was caused by a recently emerged IOL strain carrying mutations at functionally important sites. These findings highlight the importance of continued genomic surveillance to better understand viral evolution, transmission dynamics, and potential effects on virulence and immunity.

## Introduction

Chikungunya has been causing significant outbreaks in the last two decades and is now reported in over 119 countries[1]. The chikungunya virus (CHIKV) is estimated to cause 33.7 million infections annually, affecting approximately 8.4% of the population during outbreaks that last 6.2 years[2]. Over the past 20 years, the global incidence of chikungunya has risen substantially escalating from 0.28 cases per population in 2004 to 11.13 cases per 100,000 population in 2024 [3]. Large outbreaks were reported in many countries in 2025, in Latin America, South Asia, French La Reunion islands, the African continent and China[1]. Although chikungunya is an acute febrile illness, many experience debilitating joint pain with significant disability for one or two months [4]. However, as 4.7% to 78.6% have shown to progress to chronic arthritis (lasting >3 months)[5]. chikungunya is estimated to result in 1.95 million disability-adjusted life years (DALYs) and $2.8 billion in direct costs and $47.1 billion in indirect costs worldwide, between 2011 and 2020 [6]. Due to the marked rise in chikungunya cases in recent years the WHO has recommended to intensify laboratory surveillance to facilitate more effective risk communication and timely vector control strategies[1]. Due to this increasing threat of intensified chikungunya transmission, affecting a significant proportion of the global population in the tropics and subtropics, Coalition for Epidemic Preparedness Innovations (CEPI), has prioritized advancing three chikungunya vaccines, one of which is licensed and the rest are in late-phase development [7]. Therefore, it is crucial to characterize the evolution of the virus, which could adapt to increased transmissibility within vectors and persistence.

There are three main lineages (East/Central/South African, West African and Asian) of the CHIKV, which have been responsible for outbreaks during the last two decades in many countries in the tropics and sub-tropics [8]. The Indian ocean lineage (IOL), evolved from the East/Central/South African (ECSA) major lineage, and has been causing outbreaks predominantly in South Asia, Southeast Asia and the islands of the Indian ocean [8]. While the initial outbreaks of chikungunya in India during the 1960 and 1970s was due to the Asian genotype, this was replaced by the IOL arising from the ECSA genotype from outbreaks that occurred from 2005 onwards [9]. The same lineage was found to cause outbreaks in India, Bangladesh and Pakistan in recent years [10–12], although the recently reported CHIKV strains from these countries seem to have acquired different mutations from the strains that circulated prior to 2015 [13]. For instance, the CHIKV strains that most recently found to be circulating in India did not have the E1:A226V mutation, which increases transmissibility by *Aedes albopictus*, but had the E1:K211E, which increases fitness for *Aedes aegypti* [10,13]. The impact of these mutations on overall transmission of the CHIKV has not been studied. Furthermore, the virus has also acquired certain mutations such as nsP4: R82S mutation giving rise to a sub-lineage, which has previously been shown to enhance viral replication in human hosts [13]. Therefore, given the CHIKV virus is evolving and causing large outbreaks in many regions, it is important to characterize the virus causing outbreaks in different regions.

Sri Lanka experienced the first outbreak of Chikungunya between the years 2006 to 2008, which led to a 37,667 clinically suspected infections [14]. Although Chikungunya had caused outbreaks in the region, including in India from 1960s onwards, serosurveys have shown that Sri Lankans were unlikely to have been infected with the virus prior to 2006[14]. After the outbreak waned in 2008, Chikungunya cases were not reported in Sri Lanka, with recent age-stratified seroprevalence studies from Colombo, Sri Lanka showing that almost no individuals aged <16 years had evidence of prior infection before 2025[15]. However, febrile surveillance during years 2017 to 2018, has shown that in some regions in Sri Lanka, around 1% of patients presenting with a febrile illness had Chikungunya, showing that the virus had been circulating in Sri Lanka, although was not causing outbreaks [16]. Sri Lanka experienced a large outbreak due to Chikungunya, in 2025, which commenced in December 2024[17]. In this study, we characterize the molecular characteristics of CHIKV responsible for the recent large outbreak in Sri Lanka, demonstrating the presence of unique mutations leading to the emergence of a distinct Indian Ocean lineage (IOL) strain/sub-lineage.

## Methodology

### Recruitment of Patients

A total of 268 acute febrile patients presenting to the primary health-care facility of the University of Sri Jayewardenepura, Colombo, between April and September 2025 were recruited for this study. Patients with clinical symptoms suggestive of chikungunya virus (CHIKV) infection and a duration of illness of less than 10 days were enrolled following the provision of written informed consent. Ethical approval for the study was obtained from the Ethics Review Committee of the University of Sri Jayewardenepura.

### Screening for CHIKV using quantitative PCR

Viral RNA was extracted from serum samples using the MagMAX™ Viral/Pathogen Nucleic Acid Isolation Kit (Applied Biosystems™, Thermo Fisher Scientific, USA), according to the manufacturer’s protocol. All 268 samples were subsequently screened for chikungunya virus (CHIKV), dengue virus (DENV), and Zika virus (ZIKV) using a TIANLONG multiplex quantitative real-time PCR (qPCR) assay following the manufacturer’s instructions.

### Library preparation and sequencing of CHIKV

CHIKV-positive samples with cycle threshold (Ct) values <20 were selected for downstream sequencing. Complementary DNA (cDNA) synthesis was performed using LunaScript™ RT SuperMix (New England Biolabs, USA) according to the manufacturer’s instructions. Amplicon generation was carried out using a previously described tiling PCR approach specific for chikungunya virus [18]. Two primer pools (10 μM each), designated Pool A and Pool B, were used in combination with Q5® High-Fidelity DNA Polymerase (New England Biolabs, USA) and nuclease-free water. PCR amplification was performed on a QuantStudio™ 5 Real-Time PCR Instrument (Applied Biosystems, Singapore) under the following cycling conditions: an initial denaturation at 98 °C for 30 seconds, followed by 40 cycles of 98 °C for 15 seconds and 65 °C for 5 minutes, with a final hold at 4 °C.

Sequencing libraries were prepared from amplified products using the Oxford Nanopore Technologies (ONT) Rapid Barcoding Kit (SQK-RBK110.96), following the manufacturer’s protocol (RBK_9126_v110_revO_24Mar2021). Barcoded libraries were pooled and loaded onto a MinION Mk1B device (Oxford Nanopore Technologies, Oxford, United Kingdom) equipped with an R9.4 flow cell for sequencing. Real-time basecalling was performed using MinKNOW v25.05.14 (MinKNOW Core v6.5.14; Configuration v6.5.7; Bream v8.5.4) with Dorado v7.9.8 for high-accuracy base calling.

### Sequence Analysis

Raw Nanopore sequencing reads were quality-filtered using fastplong (v0.4.1)[19] in long-read mode, applying a minimum mean quality threshold of Q7. Filtered reads were subsequently used to generate sample-specific consensus sequences using a two-pass, reference-guided approach. In the first pass, reads were aligned to the chikungunya virus (CHIKV) reference genome (NC_004162.2) [20] using minimap2 (v2.30)[21]. Variant calling was performed with bcftools (v1.22)[22] using mpileup with the Oxford Nanopore-specific error model (-X ont), followed by haploid variant calling. Identified variants were applied to the reference genome to generate an initial consensus sequence.

In the second pass, reads were realigned to the first-pass consensus sequence to reduce reference bias, and variant calling was repeated to generate an updated consensus sequence. Positions with low coverage (<10×), identified using bedtools (v2.31.1)[23], were masked with ambiguous nucleotides (Ns). The resulting masked consensus sequences were used for downstream analyses.

For public data submission, only consensus sequences with >80% genome coverage relative to the reference genome (NC_004162.2), including >80% coverage across both structural and non-structural regions, were retained. Sequences meeting these completeness criteria were submitted to GenBank (NCBI) under the following accession numbers: PX661557–PX661575.

Details regarding genome recombination analysis, phylogenetic tree generation, mutational analysis, protein structural analysis and phylodynamics analyses are included in supplementary methods.

## Results

### Detection of CHIKV infection

Between April - September 2025, 100/ 268 patients presenting with clinical features suggestive of chikungunya virus (CHIKV) infection tested positive for CHIKV by multiplex quantitative real-time PCR (qPCR), while 11 patients tested positive for the dengue virus (DENV). CHIKV–DENV co-infection was detected in one patient, while no patient tested positive for the zika virus. All individuals with confirmed CHIKV infection reported fever, arthralgia, and myalgia.

Among CHIKV-positive patients, 59 were female and 41 were male. Cycle threshold (Ct) values for CHIKV ranged from 14 to 36, indicating high to low viral loads. Twenty samples with Ct values ≤20 were selected for whole-genome sequencing using Oxford Nanopore Technologies (ONT). Of these, 19 samples yielded genome coverage exceeding 80% and were included in downstream genomic and phylogenetic analyses.

### Phylogenetic analysis of the CHIKV sequences from the current outbreak

Phylogenetic analysis of CHIKV whole-genome sequences demonstrated that all Sri Lankan sequences from 2025 (n=19) clustered within the Indian Ocean Lineage (IOL) of the East/Central/South African (ECSA) genotype forming a well-supported monophyletic clade (ultrafast bootstrap support = 100%) (Figure 1). This clade was nested within a broader cluster of South Asian sequences including isolates from India (2020–2024) and Pakistan (2024) indicating regional connectivity and recent shared ancestry in transmission dynamics. In contrast, Sri Lankan sequences from the 2006–2008 outbreak (n=21) were distributed across multiple basal positions within the IOL, forming several distinct sub-clusters interspersed with sequences from diverse geographic regions.

**Figure 1:**
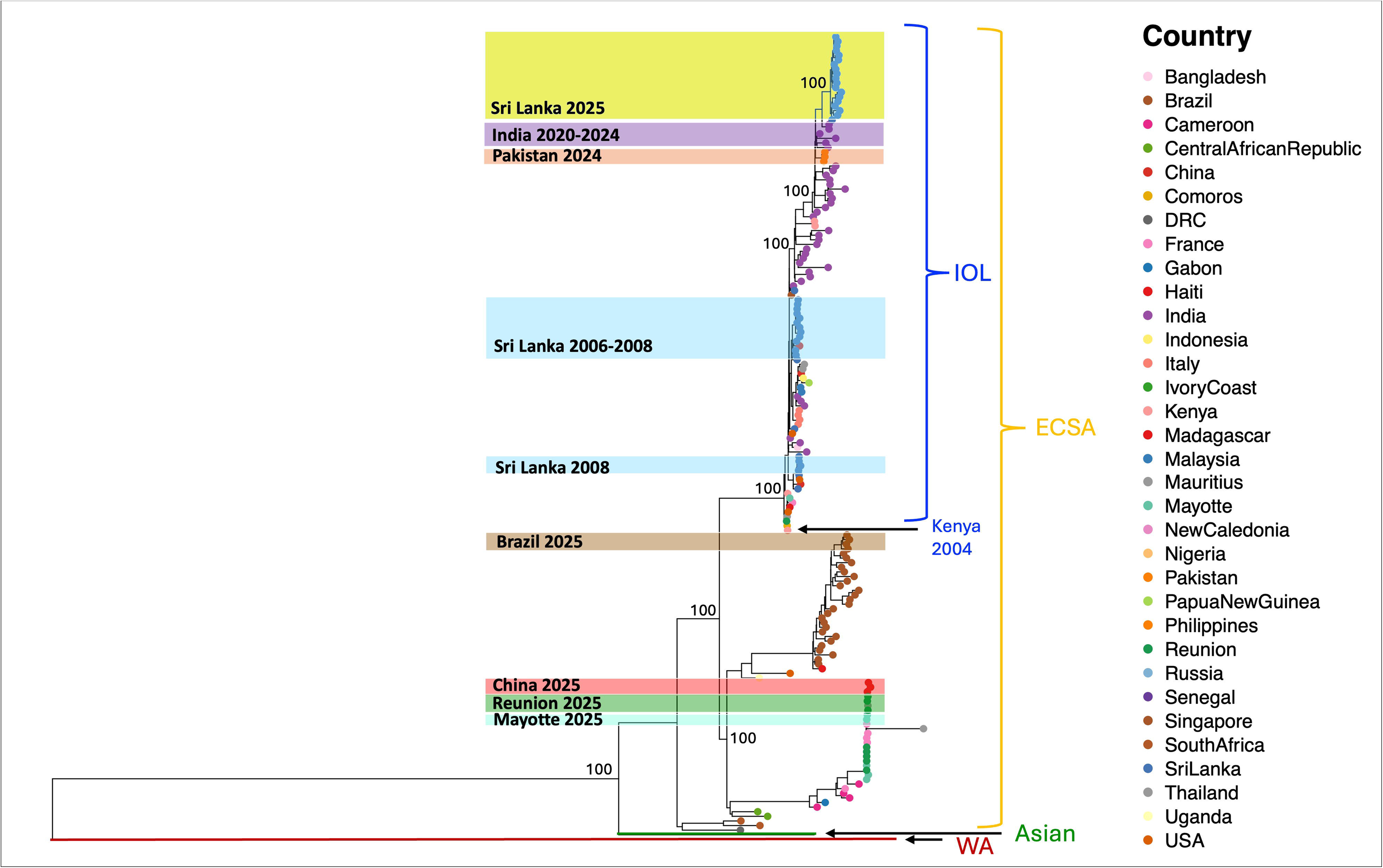
Maximum-likelihood (ML) phylogenetic tree reconstructed using complete chikungunya virus (CHIKV) genomes. Major CHIKV lineages are annotated including West African (WA), Asian, East/Central/South African (ECSA) and the Indian Ocean Lineage (IOL) with lineage boundaries indicated on the right. The tree includes Sri Lankan 2006-2008 sequences (highlighted in blue) and 2025 (highlighted in yellow) along with global reference sequences.

At the global level, the ECSA genotype exhibited a structured phylogenetic topology comprising three major groupings: (i) the Indian Ocean Lineage (IOL) which included the Sri Lanka 2025 sequences alongside isolates from India and Pakistan (ii) a distinct Brazil 2025 clade and (iii) a separate clade comprising sequences from China, Réunion, and Mayotte collected in 2025. The 2025 Sri Lankan sequences were exclusively within the IOL and did not cluster with other global lineages circulating at the same time (Figure 1). This pattern indicates that the current outbreak in Sri Lanka is likely driven by a regionally circulating lineage.

### Mutation analysis within the structural proteins (E1 and E2)

The E1 protein of the 2025 Sri Lankan sequences exhibited a distinct set of substitutions such as I55V, K211E, and D284E, which were present in all sequences within this cluster and absent in the Brazil, China, Réunion and Mayotte 2025 sequences (Figure 2). These substitutions define a distinct molecular signature of the CHIKV strains of the Sri Lankan outbreak and further support the monophyletic clustering observed in phylogenetic analysis. In contrast, sequences from Brazil 2025 exhibited a separate substitution profile, including K211T, T288I, A305T, A377V, and M407L indicating divergence from the IOL. Meanwhile, sequences from China, Réunion and Mayotte shared a different set of substitutions notably A226V along with N9S, T37I, S250P, V399I, M269V, G348E, I317V and K324R highlighting the presence of a distinct lineage within the ECSA genotype. The presence of A226V exclusively in this group further supports its separation from the Sri Lankan and South Asian clusters. The 2025 Sri Lankan sequences were uniformly characterized by I55V and K211E, both absent in the 2006 to 2008 outbreak strains (Supplementary Figure 1). In contrast, earlier sequences predominantly harbored A226V and M269V, which were not detected in 2025. These mutually exclusive patterns indicate a clear shift in the molecular profile of circulating CHIKV between the two outbreak periods.

**Figure 2:**
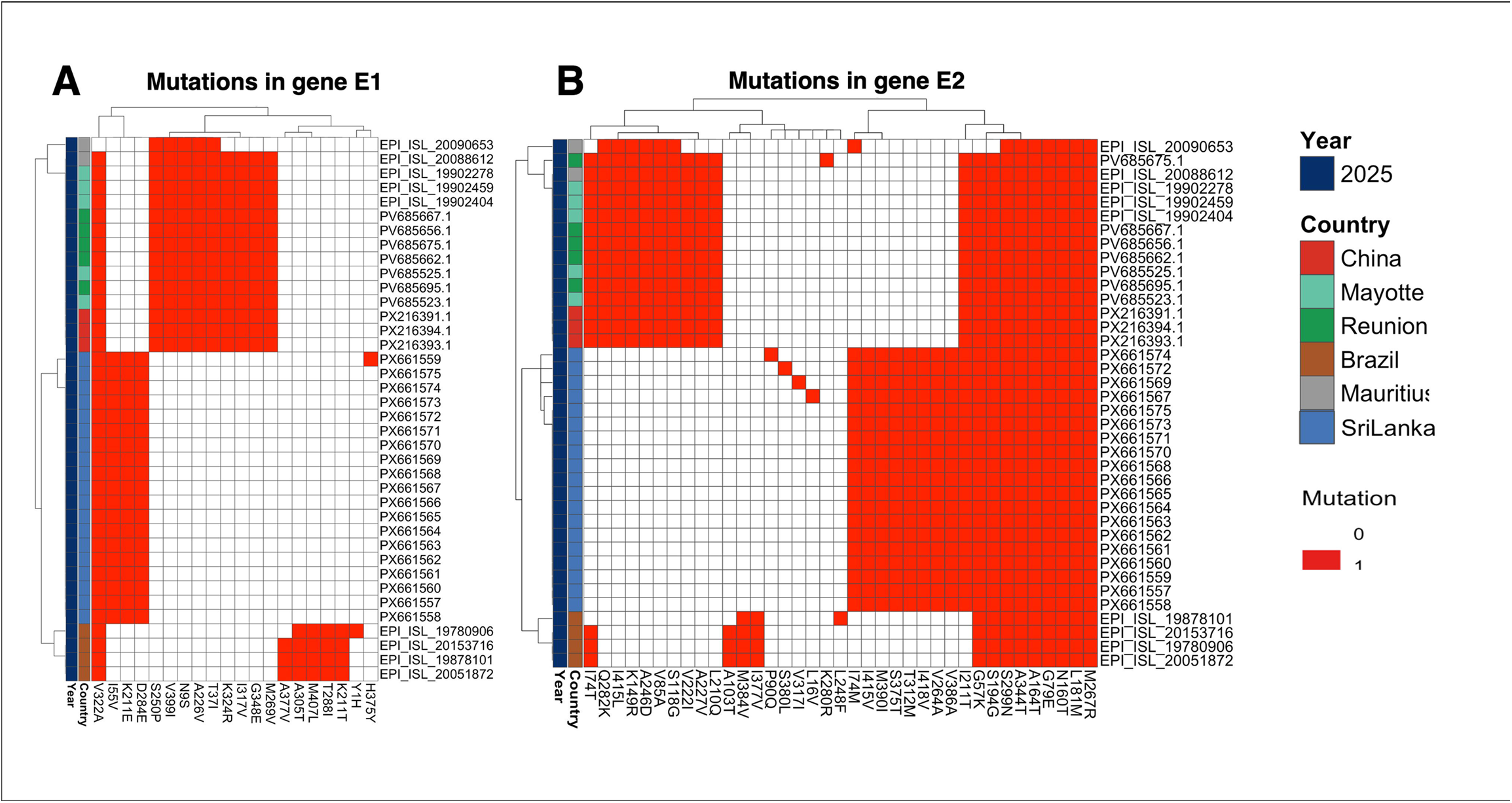
Heatmap of amino acid mutations in structural protein (E1 -E2) envelope proteins of Sri Lankan CHIKV sequences from 2025 in comparison to the CHIKV sequences circulating in year 2025 across the globe. Each row represents an individual viral genome, and each column corresponds to a specific amino acid position relative to the reference sequence (NC_004162.2). Mutations are indicated by dark red while the absence of mutation is shown in white.

The E2 protein Sri Lanka 2025 sequences carried a distinct set of substitutions, which includes V264A, M390I, I415V, I418V, I74M, S375T, T312M, and V386A. None of these mutations were detected in the Brazil or China/Réunion/Mayotte sequences. These mutations were highly conserved within the Sri Lankan cluster indicating limited intra-lineage variability and recent emergence. The 2025 CHIKV sequences from Brazil displayed harbored mutations A103T, I377V and M384V distinguishing them from both the Sri Lankan and other global lineages. In contrast, sequences from China, Réunion, and Mayotte exhibited a broader set of substitutions, including A246D, I415L, K149R, Q282K, S118G, V85A, A227V, L210Q, and V222I indicating a separate evolutionary trajectory within the ECSA genotype. Notably, the I74T substitution was shared between the Brazil and China, Réunion, and Mayotte groups but was absent in the 2025 Sri Lankan sequences, further reinforcing the distinct molecular profile of the Sri Lankan lineage.

The Sri Lanka 2025 sequences consistently carried V264A, M390I, I415V and I418V all of which were absent from the 2006–2008 sequences. These substitutions were conserved across all 2025 genomes, indicating minimal variability and recent emergence. At E2 positions 222 and 198, the 2025 sequences retained V222 and R198, whereas V222I and R198Q were seen in a subset of 2006 to 2008 sequences. Together, this defines a distinct E1 and E2 substitution profile that differentiates the 2025 outbreak from the 2006 to 2008 viruses.

### Mutation analysis within the non-structural proteins (nsp1-nsp4)

The Sri Lanka 2025 sequences were defined by the substitutions T128K, I167V, and Q488R, within the nsP1, which were absent from both Brazil and China/Réunion/Mayotte lineages (Figure 3). In contrast, Brazil sequences exhibited a lineage-specific substitution I149V, while the China/Réunion/Mayotte lineage was characterized by a distinct set of substitutions including N95S, P29S, D75E, V156I, A487T, Q488K and E498K. None of these mutations were detected in the Sri Lanka sequences.

**Figure 3:**
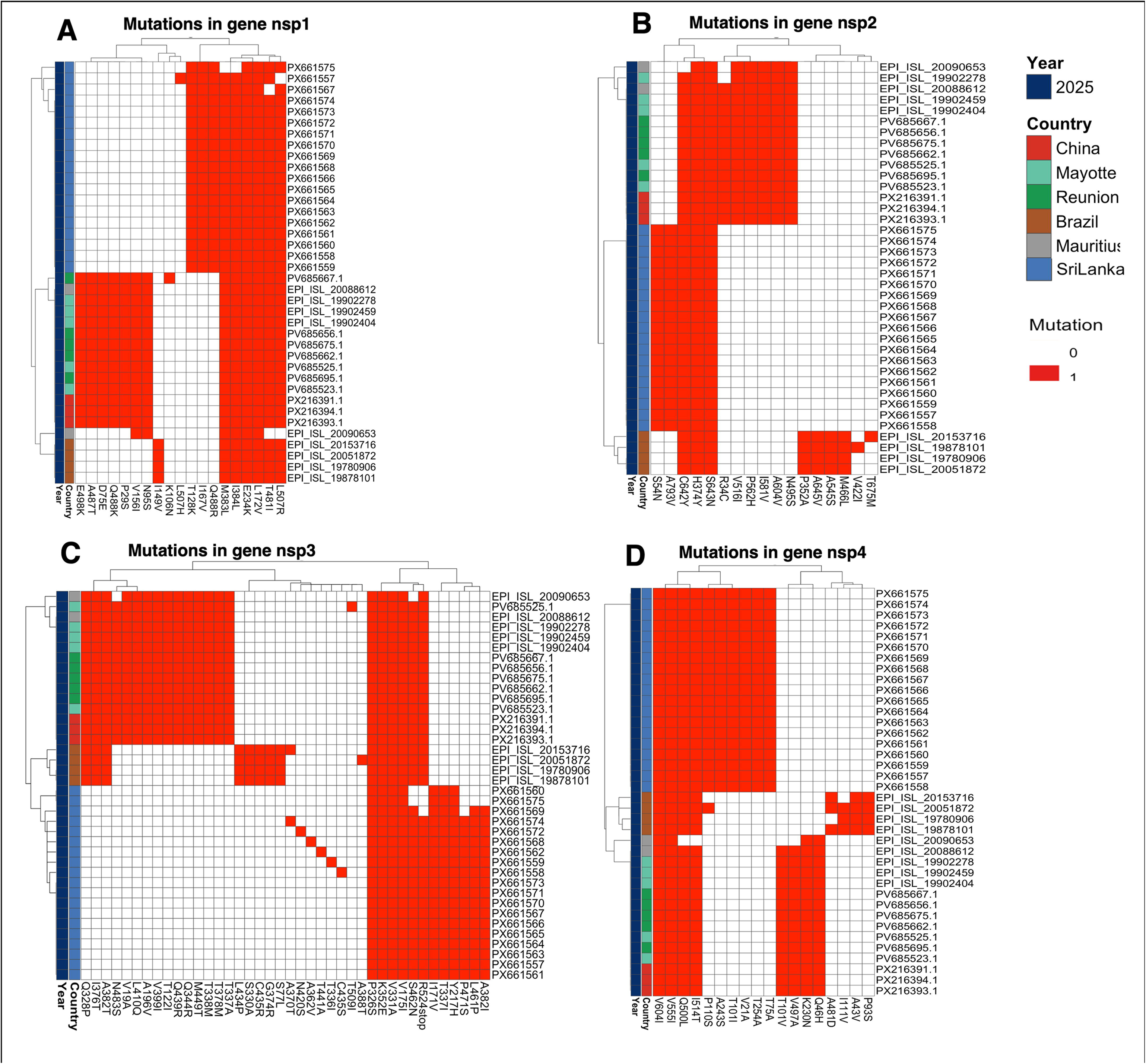
Comparative heatmap of amino acid substitutions in CHIKV non-structural proteins (nsP1–nsP4) across global and Sri Lankan sequences in year 2025. Heatmaps illustrating amino acid substitutions in the non-structural proteins of chikungunya virus (CHIKV) including (A) nsP1, (B) nsP2, (C) nsP3, and (D) nsP4. Each row represents an individual viral genome and each column corresponds to a specific amino acid position relative to the reference sequence (NC_004162.2). Mutations are indicated by dark red while the absence of mutation is shown in white.

Similar differences were seen for other non-structural proteins. Within the nsP2, Sri Lanka 2025 sequences carried S54N and A793V, which were both uniquely associated with this lineage. Sequences from Brazil showed a separate substitution profile, including P352A, M466L, A545S, and A645V whereas the China/Réunion/Mayotte lineage was characterized by R34C, N495S, V516I, I581V, P562H and A604V indicating clear divergence across lineages.

The greatest divergence was observed in nsP3, where each lineage displayed a distinct constellation of substitutions. Sri Lanka 2025 sequences were characterized by Y217H, I171V, T337I, A382I, L461P and P471S, which were all absent from other groups. Brazil sequences carried S77L, S330A, G374R, L434P and C435R, whereas the China/Réunion/Mayotte lineage exhibited a broader and more complex substitution pattern, including V19A, T122I, A196V, T337A, T338M, Q344R, T378M, V399I, L410Q, Q439R, M449T, and N483S. Notably, substitutions A382T, I376T, and Q328P were shared between Brazil and China/Réunion/Mayotte lineages but were absent from Sri Lanka sequences further highlighting the distinct evolutionary trajectory of the Sri Lankan lineage.

In nsP4, Sri Lanka 2025 sequences carried V21A, T101I, P110S, and A243S, all of which were absent from the other lineages. Brazil sequences exhibited substitutions A43V and P93S, while the China/Réunion/Mayotte lineage was defined by Q46H, K230N, T101V, and V497A, again demonstrating clear lineage-specific divergence.

Comparative analysis of non-structural proteins nsP1 to nsP4 showed a distinct mutation profile in the 2025 Sri Lankan sequences compared with those from 2006 to 2008 (Supplementary Figure 2). Several substitutions which were consistently present in 2025 CHIKV strains, were absent in earlier isolates, indicating lineage specific evolution. In nsP1, I167V was present and T376M absent, a pattern unique to 2025. The greatest divergence was seen in nsP3, with substitutions including I171V and A382I found only in 2025 sequences. In nsP4, V21A, T101I, P110S, and A243S were also exclusive to the 2025 lineage.

### Structural mapping of mutations on CHIKV envelope proteins

Structural mapping showed that several lineage defining mutations of the Sri Lankan 2025 CHIKV strain, map to regions implicated in receptor engagement or antibody recognition, including surfaces overlapping the MXRA8-binding region and known neutralizing antibody epitopes. In the E1 and E2 heterodimer (Figure 4A) and the trimeric spike complex (Figure 4B), the substitutions E1 K211E and E2 V264A were positioned within the MXRA8 receptor binding interface, indicating that key mutations in circulating strains occur at sites involved in viral entry. Additional residues, including E2 V222, were also located within the receptor contacting region, supporting the clustering of mutations at functionally relevant surfaces.

**Figure 4:**
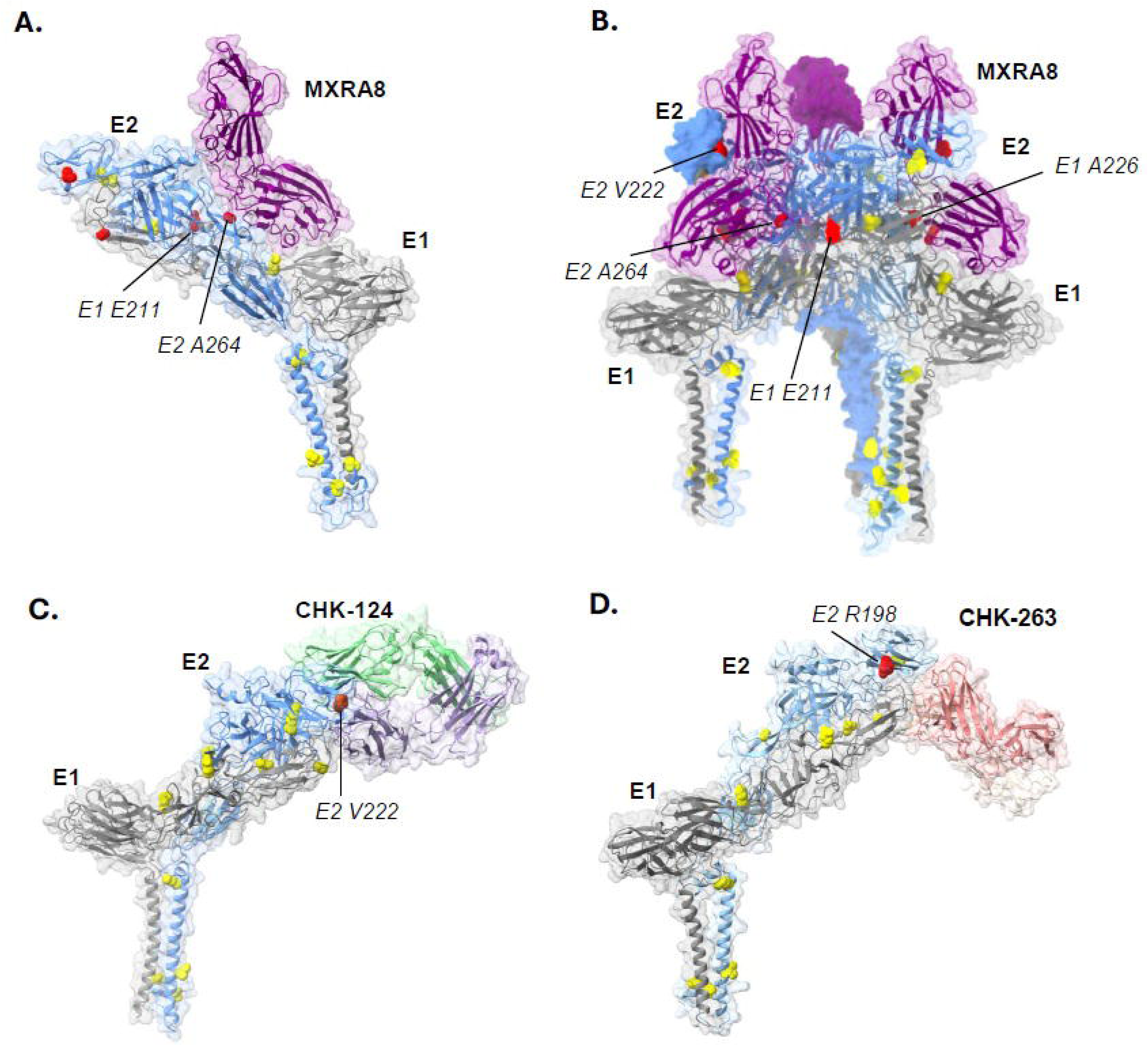
Structural mapping of amino acid substitutions in CHIKV envelope glycoproteins relative to receptor binding and neutralizing antibody epitopes. Structural representation of CHIKV envelope glycoproteins (E1–E2) highlighting the spatial distribution of amino acid substitutions identified in the 2025 Sri Lankan sequences compared to 2008 Sri Lankan sequences. (A) Docking of the host entry receptor MXRA8 onto the CHIKV E1–E2 heterodimer (B) MXRA8 binding to the trimeric spike of E1–E2 heterodimers (C) Binding of the neutralizing monoclonal antibody CHK-124 (D) Binding of the neutralizing monoclonal antibody CHK-263.

Mapping of neutralizing antibody binding sites showed that several substitutions overlap with known epitope regions. In the CHK-124 bound structure (Figure 4C), the E2 V222 substitution was located within the antibody binding interface, with additional mutations nearby. In the CHK-263 bound structure (Figure 4D), the E2 R198 substitution was positioned directly at the antibody binding site, suggesting a potential impact on antibody recognition.

### Phylodynamics analyses

Temporal signal was revealed from root-to-tip regression analysis in the program TempEst, which was demonstrated with moderate temporal structure (R2 = 0.741) (Supplementary Figure 3). While no significant evidence of recombination was detected in the CHIKV genomes used, they were subjected to the following phylodynamic analyses to explore the detailed temporal evolutionary and transmission history under more Bayesian statistical framework implemented in BEAST.

The mean rate was 8.11 × 10^−4^ nucleotide substitutions/site/year, with 95% highest posterior density (HPD) from 7.53 × 10-4 to 8.86 × 10-4. A relatively higher substitution rate inferred from BEAST can be explained by the contribution of some faster evolved strains as shown in the root-to-tip regression plot (Figure 5). The time of most recent common ancestor (tMRCA) of the clade of 2025 Sri Lankan CHIKV sequences was estimated to 2024-02 (95% HPD = 2023-08 to 2024-08), indicating spreading and unsampled circulation of those strains in Sri Lanka before the 2025 outbreak. As a sister clade, the 2025 Sri Lankan strains formed a monophyletic group with Indian strains detected in 2024, of which the tMRCA was dated to 2023-01 (95% HPD = 2022-04 to 2023-08), representing potential unsampled transmission of CHIKV in the region of India and Sri Lanka (Figure 6).

**Figure 5.**
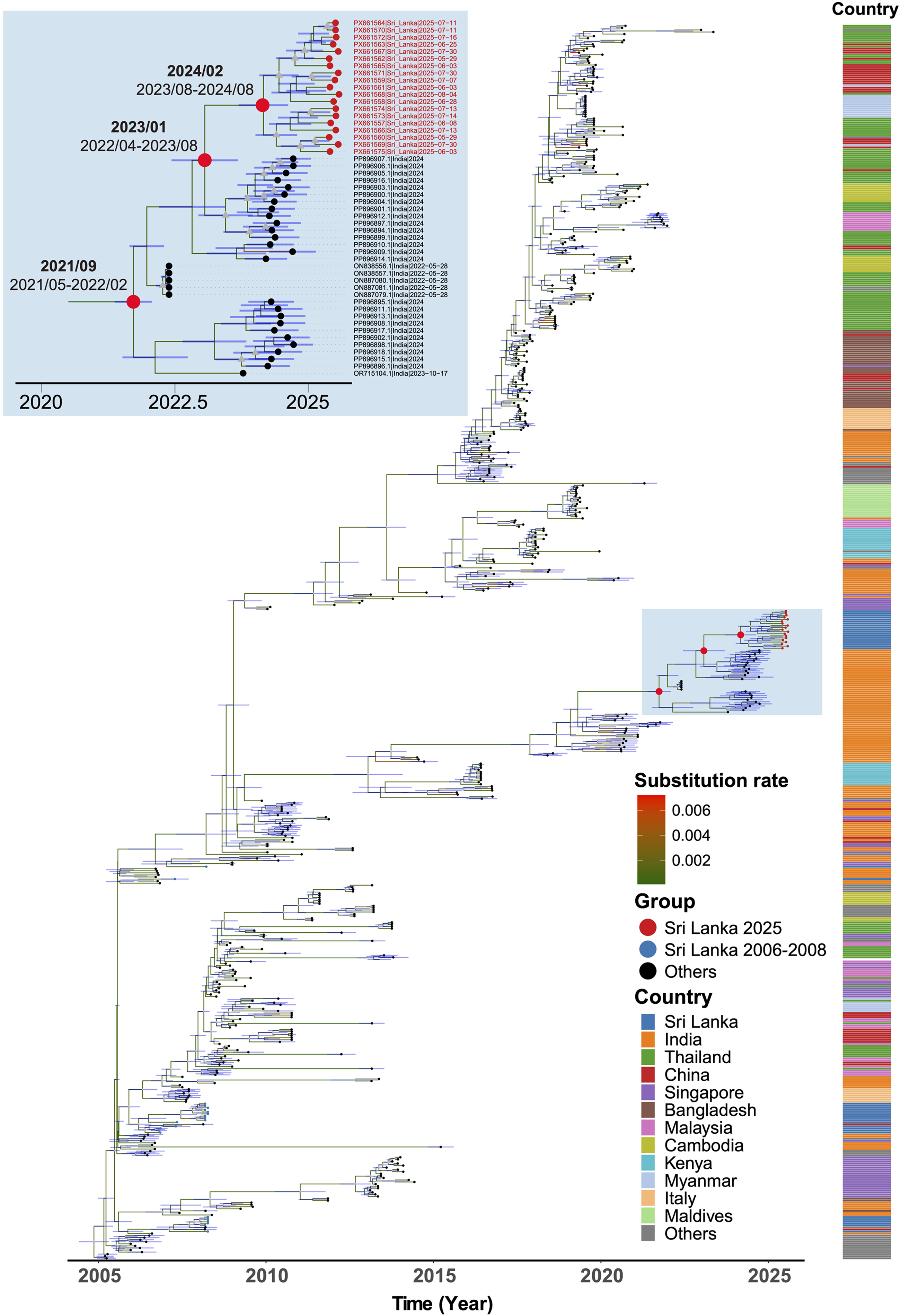
Time-scaled phylogeny of CHIKV IOL clade genomes. The maximum clade credibility tree of genome sequences (n=599) is shown. Well-supported nodes by posterior probabilities of >0.95 are indicated by grey triangles on nodes. Branches are colored by inferred nucleotide substitution rates. The tree is scaled by time of year, with node age credible intervals (95% highest posterior density [HPD]) shown on nodes. The estimated times of the most recent common ancestor of Sri Lankan 2025 sequences and its ancestral nodes are indicated on nodes with 95% HPD interval, on the top-left inset. Country of origin is annotated on the right of tips.

**Figure 6.**
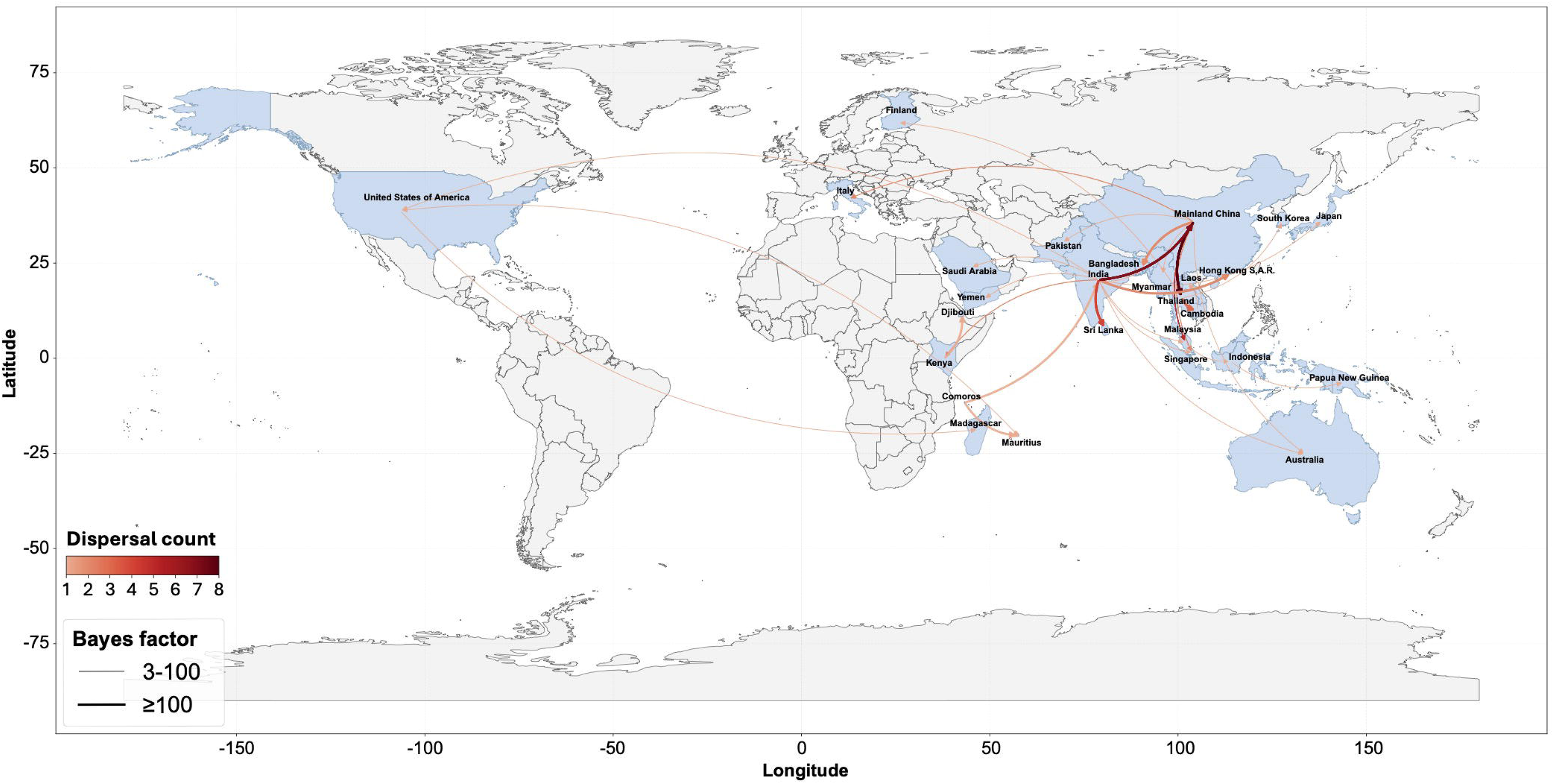
Phylogeographic dispersal map of CHIKV IOL clade genomes. Countries with the randomly sub-sampled sequences (n=172) are marked with names. Suggested transmission events are shown with arrows on the global map. The Bayes factor is indicated by the thickness of the arrow.

Bayesian phylogeographic reconstruction identified multiple statistically supported viral dispersal routes connecting Asia, Africa, Europe, and Oceania (Figure 6). Most strongly supported transmission routes occurred within Asia, indicating that this region represents the primary center of viral spread. Several long-distance dispersal events were also inferred, including movements from Asia to North America, Europe, and Africa. Within Asia, several strongly supported transmission routes (Bayes factor [BF] >= 100) were identified. Notably, strongly supported dispersal routes were inferred from China to Southeast Asia, including Thailand (BF = 215.5) and Bangladesh (BF = 141.8). A particularly strong transmission link was also observed between Thailand and Cambodia (BF = 1714.2). India appeared as a major source location in the transmission network. Strong dispersal signals were inferred from India to China (BF = 2407.7). The strongest transmission signal detected in this analysis was from India to Sri Lanka (BF = 2869.2). Multiple independent transitions were inferred along this route, including both Sri Lankan 2006-2008 and 2025 sequences, which suggested repeated viral introductions from India into Sri Lanka and highlighted the epidemiological connection between these regions.

## Discussion

Sri Lanka faced one of the largest outbreaks of Chikungunya in 2025 after 16 years, which commenced during December 2024. We found that the CHIKV causing this outbreak was significantly different to the CHIKV strains that circulated between 2006 to 2008, but similar to the CHIKV strains that circulated in India, Pakistan and Bangladesh in 2024 [24–26]. Despite CHIKV circulating in Sri Lanka during the last 16 years and causing sporadic infection [16], no outbreaks were reported. CHIKV not only suddenly emerged causing outbreaks in the South Asian region in 2024, but caused outbreaks in La Réunion islands, many countries in Latin America and in China[1]. Interestingly, although the CHIKV causing outbreaks in South Asia were within the same lineage, the CHIKV causing outbreaks in China, La Réunion and Brazil were of a different clade[27,28]. While the CHIKV from China, La Réunion and Mayotte belonged to the same lineage, the CHIKV strains in Brazil were different to the South Asian and Chinese clades[28]. While the reasons for the simultaneous emergence of multiple CHIKV lineages causing outbreaks in different regions remain unclear, these findings highlight the growing global threat posed by mosquito borne viral infections.

The currently circulating Sri Lankan CHIKV strain contained multiple mutations in the E1 and E2 structural proteins as well as in the non-structural proteins, sharing features with other South Asian strains while also showing unique mutations. The I55V, K211E, and D284E mutations within E1 define a distinct molecular signature of the CHIKV strains of the Sri Lankan and South Asian strains, which were absent from the strains from Brazil and China, La Réunion and Mayotte clusters. While China, La Réunion and Mayotte CHIKV strains contained the A226V mutation, which has been associated with *Aedes albopictus* transmission efficiency, was absent in all Sri Lankan and South Asian 2025 sequences [13]. Instead, the Sri Lankan and other South Asian sequences carried the E1:K211E and E2: V264A, which result in enhanced viral fitness within *Aedes aegypti* [29]. Therefore, these mutations could lead to higher transmission rates in urban areas, where the predominant *Aedes* specifies is *Aedes aegypti*. The emergence of the E1:K211E/E2:V264A sub lineage of the IOL, has also been responsible for the large Chikungunya outbreak reported in Malaysia in 2021 [30]. Similar mutations were also present in the Sri Lankan/South Asian CHIKV lineages, which were absent in the lineages from Brazil and China, further showing that the South Asian lineages are quite distinct from the CHIKV lineages causing outbreaks in other regions in the world.

Although the significance of the mutations of the South Asian strains in the E1 and E2 protein are not clear, the substitutions E1 K211E and E2 V264A are located within the MXRA8 receptor binding interface. The MXRA8 receptor is important for virus attachment and internalization of the CHIKV, and antibodies blocking these receptors have shown to prevent infection of cells[31]. Therefore, it would be important to find out if two key mutations within the receptor is associated with enhanced infection rates of cells. In addition to above mutations, the E2 R198 substitution was shown to be positioned within the key neutralizing antibody binding site. Although it has been shown that individuals infected with CHIKV strains have long lasting immunity[32] and cross protect against other CHIKV strains (Asian genotypes vs ESCA strains) still completely neutralized the other virus strains [33], it would be important to determine if these mutations have any influence on neutralizing ability.

Apart from the mutations detected with the envelope protein, many mutations were detected within the non-structural proteins such as nsP1:I167V, nsP3:I171V, nsP3:A382I and nsP4:V21A. Although the significance of mutations, which are also seen in the most recent circulating CHIKV strains in South Asia are unknown, the non-structural proteins of CHIKV are unknown to interact with host proteins and have also shown to be important in immune evasion [34,35]. Again, as observed with the envelope proteins, the South Asian strains had a significantly different mutation profile compared to strains from Brazil, China, La Réunion and Mayotte CHIKV strains. The greatest divergence was seen in the nsP3 protein. The nsP3 protein is essential for virus replication and also inhibits many down-stream immune signaling pathways[34,36]. Therefore, mutations within this region may affect viral fitness and virulence.

The global phylogeographic analysis suggests that the CHIKV strain responsible for the 2025 outbreak in Sri Lanka likely originated from viruses circulating in India. India appeared to serve as a major source of transmission in the region, with dissemination to countries such as Saudi Arabia, Sri Lanka, Japan, Hong Kong, and Mainland China. The results also suggest that China functions as another significant source for virus transmission to neighboring countries, including Thailand and Malaysia. Although the Bayesian phylogeographic method used in this study accounts for statistical uncertainty, and variability in dissemination and evolutionary rates, the findings may still be influenced by sampling biases across different geographical regions. Expanding sequencing efforts to include more representative samples from diverse regions will further strengthen the confidence in addressing this issue.

In summary, we characterized the CHIKV strains responsible for the large outbreak in Sri Lanka in 2025. These strains are similar to those currently circulating in South Asia, but distinct from lineages causing outbreaks in Brazil, China, La Réunion, and Mayotte. The Sri Lankan lineage is defined by several key mutations, most shared with regional strains but some unique. Notably, mutations in the E1 and E2 proteins were located within the MXRA8 receptor binding domain and key neutralizing antibody binding regions. The factors driving the resurgence of multiple CHIKV lineages across different regions, and whether viral evolution has contributed to increased virulence, require further investigation. However, the limited sequencing fraction due to selection of samples with low Ct values and sampling limited to an urban cohort may not reflect full viral diversity, and the functional effects of identified mutations remain to be experimentally validated.

## Supporting information

Suppmentary data

## Data availability statement

The data that support the findings of this study are openly available in the NCBI GenBank repository under accession numbers PX661557–PX661575, corresponding to the 19 newly generated CHIKV genomes from Sri Lanka. Metadata associated with these sequences are provided in Supplementary Table 1. Publicly available chikungunya virus genome sequences used for comparative phylogenetic, mutational, and phylodynamic analyses were obtained from GenBank and GISAID and are listed in Supplementary Tables 2–5. GISAID acknowledgment information is available under EPI_SET_260422gv (DOI: https://doi.org/10.55876/gis8.260422gv). Additional supporting data are available from corresponding author upon request.

## Data deposition

Newly generated chikungunya virus (CHIKV) consensus genome sequences have been deposited in GenBank under accession numbers PX661557–PX661575. Associated metadata and sequencing statistics are provided in Supplementary Table 1. Raw Oxford Nanopore sequencing reads and additional supporting data are available from the corresponding author upon request.

