## Supplementary material for "Genomic characterization of chikungunya virus during the large 2025 outbreak in Sri Lanka reveals emergence of a distinct Indian Ocean lineage strain": Suppmentary data

**Supplementary Files**

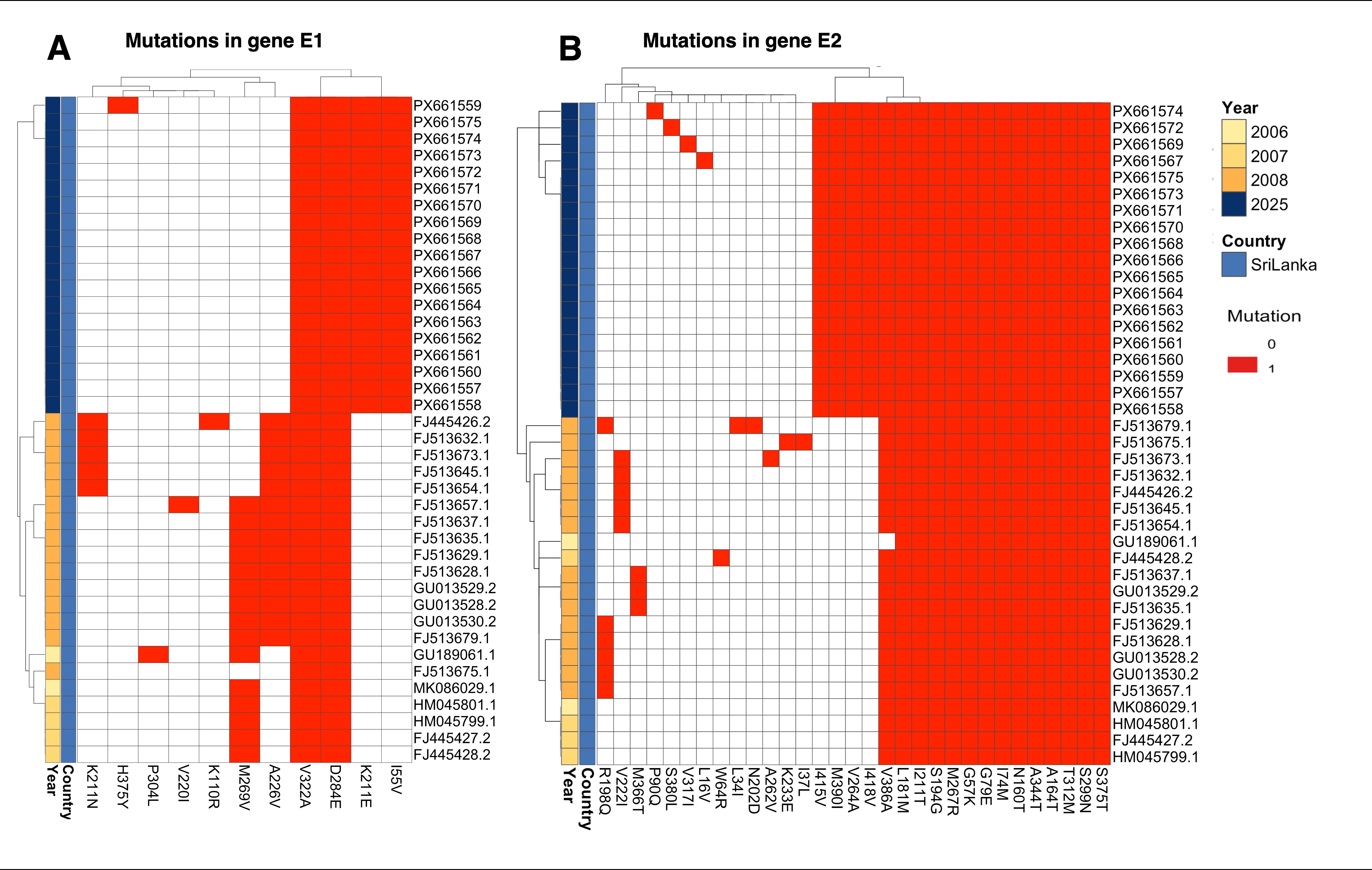

**Supplementary Figure 1: Comparative heatmap illustrating amino acid substitutions in CHIKV structural glycoproteins (E1 and E2) between sequences from the 2006–2008 outbreak and those from the 2025 outbreak in Sri Lanka**. Each row represents an individual viral genome and columns indicate amino acid substitutions relative to the reference genome (NC_004162.2). Dark red cells indicate the presence of a mutation at the corresponding position.

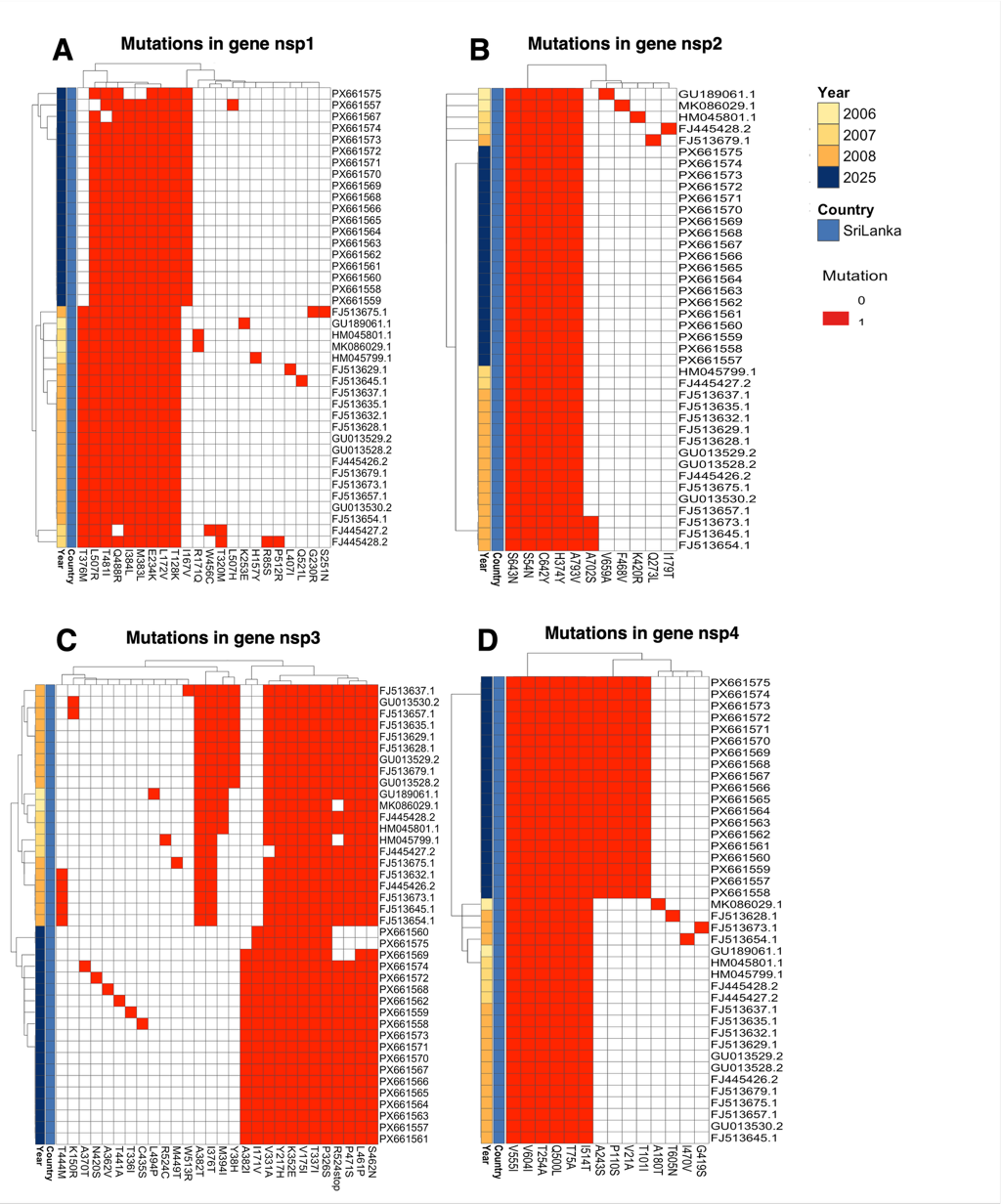

**Supplementary Figure 2. Comparative mutation profiles of CHIKV non-structural proteins between Sri Lankan outbreaks (2006–2008 vs 2025).** Heatmaps illustrating amino acid substitutions in the non-structural proteins of chikungunya virus (CHIKV), including (A) nsP1, (B) nsP2, (C) nsP3, and (D) nsP4, comparing sequences from the 2006–2008 outbreak and the 2025 outbreak in Sri Lanka. Each row represents an individual viral genome, and each column corresponds to a specific amino acid position relative to the reference sequence (NC_004162.2).

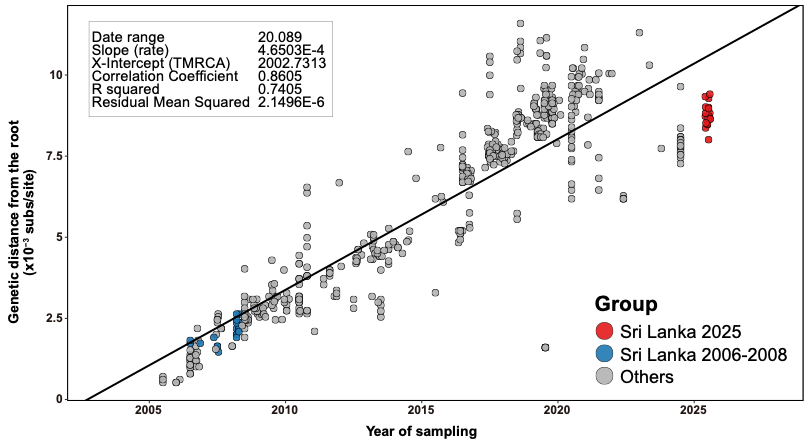

**Supplementary Figure 3.** TempEst regression for the Chikungunya virus IOL clade. Data points were color coded according to colors in Fig 5. Each point represents a genome, with the x-axis showing sampling date (in decimal years) and the y-axis representing genetic divergence from the tree root.

**Supplementary Table 1. Metadata and sequencing statistics for CHIKV genomes generated in this study (Sri Lanka, 2025)**

| **Genbank ID** | **Same_ID** | **Collection Date** | **Area** | **ct** | **raw reads** | **trimmed reads** | **Mean depth** | **Length** | **Ns** |
| --- | --- | --- | --- | --- | --- | --- | --- | --- | --- |
| PX661557 | barcode73 | 2025-06-08 | Hokandara | 20 | 133913 | 128903 | 4159.32 | 11826 | 125 |
| PX661558 | barcode74 | 2025-06-28 | Habarakada | 18 | 104150 | 103670 | 3827.38 | 11826 | 126 |
| PX661559 | barcode75 | 2025-07-07 | Kottawa | 16 | 146462 | 142364 | 5649.01 | 11826 | 125 |
| PX661560 | barcode76 | 2025-05-29 | Battaramulla | 20 | 74511 | 72904 | 1369.2 | 11826 | 995 |
| PX661561 | barcode77 | 2025-06-03 | Mattegoda | 17 | 163118 | 156271 | 6430.54 | 11826 | 125 |
| PX661562 | barcode78 | 2025-05-29 | Athurugiriya | 18 | 157000 | 156749 | 5992.91 | 11826 | 125 |
| PX661563 | barcode79 | 2025-06-25 | Kottawa | 20 | 290807 | 277176 | 12494.6 | 11826 | 124 |
| PX661564 | barcode80 | 2025-07-11 | Hokandara | 20 | 186711 | 181933 | 7343.89 | 11826 | 125 |
| PX661565 | barcode81 | 2025-06-03 | Kotte | 18 | 94074 | 94008 | 3298.78 | 11826 | 126 |
| PX661575 | barcode82 | 2025-06-03 | Kaduwela | 19 | 63362 | 62485 | 885.001 | 11826 | 1432 |
| PX661566 | barcode83 | 2025-07-13 | Athurugiriya | 14 | 155391 | 150002 | 5971.72 | 11826 | 125 |
| PX661567 | barcode84 | 2025-07-30 | Pannipitiya | 21 | 201050 | 193384 | 7669.81 | 11826 | 130 |
| PX661568 | barcode85 | 2025-08-04 | Rukmale | 18 | 194553 | 187095 | 8439.25 | 11826 | 125 |
| PX661569 | barcode86 | 2025-07-30 | Ragama | 14 | 80476 | 79508 | 1651.68 | 11826 | 356 |
| PX661570 | barcode87 | 2025-07-11 | Hokandara | 16 | 168314 | 161574 | 6727.07 | 11826 | 125 |
| PX661571 | barcode88 | 2025-07-30 | Malabe | 20 | 198144 | 192243 | 7547.55 | 11826 | 125 |
| PX661572 | barcode89 | 2025-07-16 | Malabe | 20 | 119040 | 117710 | 4803.29 | 11826 | 125 |
| PX661573 | barcode91 | 2025-07-14 | Malabe | 16 | 118630 | 118523 | 5445.85 | 11826 | 131 |
| PX661574 | barcode92 | 2025-07-13 | Malabe | 18 | 188245 | 180265 | 8328.98 | 11826 | 144 |

**Supplementary Table 2. Public CHIKV genomes included in phylogenetic analyses (n = 225)**

| ID | Country | Collection_Date | Database_Type |
| --- | --- | --- | --- |
| HQ456254.1 | Kenya | 2004-10-01 | GenBank |
| HQ456255.1 | Kenya | 2004-07-01 | GenBank |
| HQ456251.1 | Comoros | 2005-03-01 | GenBank |
| PX216394.1 | China | 2025-07-31 | GenBank |
| PX216393.1 | China | 2025-07-27 | GenBank |
| PX216391.1 | China | 2025-07-24 | GenBank |
| PV685523.1 | Mayotte | 2025-04-11 | GenBank |
| PV685695.1 | Reunion | 2025-04-02 | GenBank |
| PV685525.1 | Mayotte | 2025-03-29 | GenBank |
| PV685662.1 | Reunion | 2025-03-20 | GenBank |
| PV685675.1 | Reunion | 2025-03-13 | GenBank |
| PV685656.1 | Reunion | 2025-02-25 | GenBank |
| PV685667.1 | Reunion | 2025-02-10 | GenBank |
| PV685637.1 | Reunion | 2024-12-18 | GenBank |
| PV685641.1 | Reunion | 2024-12-18 | GenBank |
| PV685636.1 | Reunion | 2024-12-12 | GenBank |
| PP798381.1 | China | 2024-01-22 | GenBank |
| PP896896.1 | India | 2024-01-01 | GenBank |
| PP896898.1 | India | 2024-01-01 | GenBank |
| PP896903.1 | India | 2024-01-01 | GenBank |
| PP896906.1 | India | 2024-01-01 | GenBank |
| PP896909.1 | India | 2024-01-01 | GenBank |
| PP896911.1 | India | 2024-01-01 | GenBank |
| PP896913.1 | India | 2024-01-01 | GenBank |
| OR715104.1 | India | 2023-10-17 | GenBank |
| PP319439.1 | Haiti | 2021-04-15 | GenBank |
| OQ605454.1 | India | 2021-01-01 | GenBank |
| OL898675.1 | Brazil | 2020-12-26 | GenBank |
| OL898669.1 | Brazil | 2020-12-17 | GenBank |
| OL898714.1 | Brazil | 2020-11-20 | GenBank |
| OL898663.1 | Brazil | 2020-09-05 | GenBank |
| OQ605436.1 | India | 2020-01-01 | GenBank |
| OQ605439.1 | India | 2020-01-01 | GenBank |
| OQ605441.1 | India | 2020-01-01 | GenBank |
| OQ605450.1 | India | 2020-01-01 | GenBank |
| OK316989.1 | China | 2019-11-11 | GenBank |
| OK316990.1 | China | 2019-11-11 | GenBank |
| MT666071.1 | Cameroon | 2018-09-17 | GenBank |
| MT666073.1 | Cameroon | 2018-06-28 | GenBank |
| PP896920.1 | India | 2018-01-01 | GenBank |
| MW281311.1 | France | 2017-09-12 | GenBank |
| MH823664.1 | Brazil | 2017-03-16 | GenBank |
| MH823665.1 | Brazil | 2017-03-16 | GenBank |
| MT666072.1 | Cameroon | 2016-10-20 | GenBank |
| MK473626.1 | India | 2016-10-06 | GenBank |
| MK518340.1 | India | 2016-09-28 | GenBank |
| OQ148637.1 | Brazil | 2016-06-15 | GenBank |
| MH423804.1 | Kenya | 2016-06-01 | GenBank |
| MH423803.1 | Kenya | 2016-05-31 | GenBank |
| LC259091.1 | Indonesia | 2015-05-07 | GenBank |
| PV089294.1 | Brazil | 2014-08-07 | GenBank |
| KM673291.1 | Indonesia | 2013-01-01 | GenBank |
| MF773569.1 | PapuaNewGuinea | 2013-01-01 | GenBank |
| LC259085.1 | Indonesia | 2012-10-15 | GenBank |
| ON262791.1 | Thailand | 2012-01-01 | GenBank |
| MG664851.1 | China | 2012-01-01 | GenBank |
| KJ679578.1 | India | 2011-12-21 | GenBank |
| HE806461.1 | NewCaledonia | 2011-02-28 | GenBank |
| MH124574.1 | India | 2010-01-01 | GenBank |
| MH124575.1 | India | 2010-01-01 | GenBank |
| MH124576.1 | India | 2010-01-01 | GenBank |
| MH124578.1 | India | 2010-01-01 | GenBank |
| KC862329.1 | Indonesia | 2010-01-01 | GenBank |
| MH670649.1 | China | 2009-11-19 | GenBank |
| GU301780.1 | Thailand | 2008-10-21 | GenBank |
| GU013530.2 | SriLanka | 2008-04-01 | GenBank |
| FJ513645.1 | SriLanka | 2008-04-01 | GenBank |
| FJ513654.1 | SriLanka | 2008-04-01 | GenBank |
| FJ513657.1 | SriLanka | 2008-04-01 | GenBank |
| FJ513673.1 | SriLanka | 2008-04-01 | GenBank |
| FJ513675.1 | SriLanka | 2008-04-01 | GenBank |
| FJ513679.1 | SriLanka | 2008-04-01 | GenBank |
| FJ445426.2 | SriLanka | 2008-04-01 | GenBank |
| GU013528.2 | SriLanka | 2008-03-01 | GenBank |
| GU013529.2 | SriLanka | 2008-03-01 | GenBank |
| FJ513628.1 | SriLanka | 2008-03-01 | GenBank |
| FJ513629.1 | SriLanka | 2008-03-01 | GenBank |
| FJ513632.1 | SriLanka | 2008-03-01 | GenBank |
| FJ513635.1 | SriLanka | 2008-03-01 | GenBank |
| FJ513637.1 | SriLanka | 2008-03-01 | GenBank |
| KY575570.1 | USA | 2008-01-01 | GenBank |
| JN558835.1 | India | 2008-01-01 | GenBank |
| GU199350.1 | China | 2008-01-01 | GenBank |
| GU199351.1 | China | 2008-01-01 | GenBank |
| GU199352.1 | China | 2008-01-01 | GenBank |
| FJ807898.1 | Bangladesh | 2008-01-01 | GenBank |
| FJ807899.1 | Malaysia | 2008-01-01 | GenBank |
| FN295485.3 | Malaysia | 2008-01-01 | GenBank |
| MK120202.1 | Italy | 2007-08-25 | GenBank |
| MK120201.1 | Italy | 2007-08-20 | GenBank |
| GQ428212.1 | India | 2007-07-12 | GenBank |
| FJ445427.2 | SriLanka | 2007-07-01 | GenBank |
| FJ000069.1 | India | 2007-06-01 | GenBank |
| FJ445428.2 | SriLanka | 2007-05-01 | GenBank |
| KX262989.1 | Italy | 2007-01-01 | GenBank |
| KX262993.1 | Italy | 2007-01-01 | GenBank |
| KP003812.1 | Gabon | 2007-01-01 | GenBank |
| HM045799.1 | SriLanka | 2007-01-01 | GenBank |
| HM045801.1 | SriLanka | 2007-01-01 | GenBank |
| MK086029.1 | SriLanka | 2006-11-14 | GenBank |
| EU564335.1 | India | 2006-10-31 | GenBank |
| GQ428211.1 | India | 2006-10-07 | GenBank |
| FN295483.3 | Malaysia | 2006-03-01 | GenBank |
| FN295484.2 | Malaysia | 2006-03-01 | GenBank |
| EU564334.1 | Mauritius | 2006-02-14 | GenBank |
| KY575571.1 | USA | 2006-01-01 | GenBank |
| KX262996.1 | Cameroon | 2006-01-01 | GenBank |
| KP003807.1 | France | 2006-01-01 | GenBank |
| KP003808.1 | Madagascar | 2006-01-01 | GenBank |
| KP003809.1 | Mayotte | 2006-01-01 | GenBank |
| HM045794.1 | USA | 2006-01-01 | GenBank |
| GU189061.1 | SriLanka | 2006-01-01 | GenBank |
| FJ807896.1 | Singapore | 2006-01-01 | GenBank |
| EU703759.1 | Malaysia | 2006-01-01 | GenBank |
| EU703761.1 | Malaysia | 2006-01-01 | GenBank |
| EU703762.1 | Malaysia | 2006-01-01 | GenBank |
| FR717336.1 | Reunion | 2005-12-26 | GenBank |
| KX262987.1 | Thailand | 1996-01-01 | GenBank |
| KY575574.1 | USA | 1995-01-01 | GenBank |
| HM045787.1 | Thailand | 1995-01-01 | GenBank |
| KX262988.1 | Thailand | 1988-01-01 | GenBank |
| HM045789.1 | Thailand | 1988-01-01 | GenBank |
| HM045797.1 | Indonesia | 1985-01-01 | GenBank |
| HM045800.1 | Philippines | 1985-01-01 | GenBank |
| KY038947.2 | CentralAfricanRepublic | 1983-12-01 | GenBank |
| HM045791.1 | Indonesia | 1983-01-01 | GenBank |
| HM045812.1 | Uganda | 1982-01-01 | GenBank |
| HM045822.1 | CentralAfricanRepublic | 1978-10-01 | GenBank |
| HM045808.1 | Thailand | 1978-01-01 | GenBank |
| HM045795.1 | SouthAfrica | 1976-01-01 | GenBank |
| HM045814.1 | Thailand | 1975-01-01 | GenBank |
| HM045788.1 | India | 1973-01-01 | GenBank |
| HM045803.1 | India | 1963-11-06 | GenBank |
| HM045813.1 | India | 1963-11-06 | GenBank |
| HM045809.1 | DRC | 1960-01-01 | GenBank |
| LC259082.1 | Thailand | 1958-01-01 | GenBank |
| HM045810.1 | Thailand | 1958-01-01 | GenBank |
| HM045792.1 | SouthAfrica | 1956-04-01 | GenBank |
| EPI_ISL_17456004 | WA_Senegal | 1966-11-23 | GISAID |
| EPI_ISL_17456015 | WA_Nigeria | 1964-07-07 | GISAID |
| EPI_ISL_18498015 | WA_Senegal | 2023-08-25 | GISAID |
| EPI_ISL_18498016 | WA_Senegal | 2023-08-24 | GISAID |
| EPI_ISL_18498019 | WA_Senegal | 2023-08-21 | GISAID |
| EPI_ISL_18498021 | WA_Senegal | 2023-08-24 | GISAID |
| EPI_ISL_18498026 | WA_Senegal | 2023-08-21 | GISAID |
| EPI_ISL_18498032 | WA_Senegal | 2023-08-18 | GISAID |
| EPI_ISL_18498035 | WA_Senegal | 2023-08-17 | GISAID |
| EPI_ISL_18498036 | WA_Senegal | 2023-08-23 | GISAID |
| EPI_ISL_18498037 | WA_Senegal | 2023-08-21 | GISAID |
| EPI_ISL_19557244 | WA_France | 2024-07-19 | GISAID |
| EPI_ISL_19557246 | WA_IvoryCoast | 2024-07-07 | GISAID |
| EPI_ISL_19557247 | WA_IvoryCoast | 2024-05-25 | GISAID |
| EPI_ISL_19557249 | WA_IvoryCoast | 2023-06-21 | GISAID |
| EPI_ISL_19557251 | WA_IvoryCoast | 2024-07-16 | GISAID |
| EPI_ISL_19557253 | WA_IvoryCoast | 2023-08-10 | GISAID |
| EPI_ISL_19557255 | WA_IvoryCoast | 2024-07-19 | GISAID |
| EPI_ISL_20146493 | WA_Senegal | 2023-08-24 | GISAID |
| EPI_ISL_17455726 | Asian_Russia | 2013-09-24 | GISAID |
| EPI_ISL_19085665 | Asian_Indonesia | 2024-01-20 | GISAID |
| EPI_ISL_17461544 | Asian_Indonesia | 2016-11-21 | GISAID |
| EPI_ISL_17461545 | Asian_Indonesia | 2016-11-24 | GISAID |
| EPI_ISL_20136536 | Asian_China | 2025-08-08 | GISAID |
| EPI_ISL_19650208 | Pakistan | 2024-11-28 | GISAID |
| EPI_ISL_19650210 | Pakistan | 2024-11-21 | GISAID |
| EPI_ISL_19902404 | Mayotte | 2025-03-22 | GISAID |
| EPI_ISL_19902459 | Mayotte | 2025-04-12 | GISAID |
| EPI_ISL_17459258 | India | 2013-03-06 | GISAID |
| EPI_ISL_17459260 | India | 2009-04-08 | GISAID |
| EPI_ISL_17459261 | India | 2012-08-03 | GISAID |
| EPI_ISL_17459262 | India | 2012-08-03 | GISAID |
| EPI_ISL_19372509 | Brazil | 2019-06-12 | GISAID |
| EPI_ISL_17460418 | Brazil | 2019-03-08 | GISAID |
| EPI_ISL_17673126 | Brazil | 2018-04-05 | GISAID |
| EPI_ISL_17954081 | Brazil | 2015-07-22 | GISAID |
| EPI_ISL_17673142 | Brazil | 2018-05-23 | GISAID |
| EPI_ISL_17954035 | Brazil | 2015-07-17 | GISAID |
| EPI_ISL_19780906 | Brazil | 2025-02-11 | GISAID |
| EPI_ISL_20075230 | France | 2025-03-13 | GISAID |
| EPI_ISL_20075236 | France | 2025-04-03 | GISAID |
| EPI_ISL_20075239 | France | 2025-03-25 | GISAID |
| EPI_ISL_20075241 | France | 2024-11-26 | GISAID |
| EPI_ISL_20217856 | Brazil | 2024-02-08 | GISAID |
| EPI_ISL_19902278 | Mayotte | 2025-03-20 | GISAID |
| EPI_ISL_17458770 | India | 2015-09-22 | GISAID |
| EPI_ISL_19372750 | India | 2021-01-04 | GISAID |
| EPI_ISL_19372751 | India | 2021-02-06 | GISAID |
| EPI_ISL_17458785 | India | 2014-07-23 | GISAID |
| EPI_ISL_17458787 | India | 2015-09-11 | GISAID |
| EPI_ISL_17459256 | India | 2010-07-16 | GISAID |
| EPI_ISL_19116421 | Brazil | 2024-01-30 | GISAID |
| EPI_ISL_19878101 | Brazil | 2025-01-30 | GISAID |
| EPI_ISL_20088612 | Mauritius | 2025-03-30 | GISAID |
| EPI_ISL_17456929 | Brazil | 2016-10-13 | GISAID |
| EPI_ISL_18553691 | Brazil | 2023-06-10 | GISAID |
| EPI_ISL_19429570 | Pakistan | 2024-06-09 | GISAID |
| EPI_ISL_20090653 | Mauritius | 2025-04-16 | GISAID |
| EPI_ISL_17954384 | Brazil | 2021-09-30 | GISAID |
| EPI_ISL_19725182 | Brazil | 2024-12-02 | GISAID |
| EPI_ISL_17954404 | Brazil | 2021-03-03 | GISAID |
| EPI_ISL_18220053 | Brazil | 2023-05-09 | GISAID |
| EPI_ISL_17455903 | India | 2019-08-28 | GISAID |
| EPI_ISL_19007928 | Brazil | 2017-03-22 | GISAID |
| EPI_ISL_19007948 | Brazil | 2016-04-28 | GISAID |
| EPI_ISL_17458379 | Brazil | 2016-03-16 | GISAID |
| EPI_ISL_20051872 | Brazil | 2025-04-22 | GISAID |
| EPI_ISL_20153716 | Brazil | 2025-03-07 | GISAID |
| PX661557 | SriLanka | 2025-06-08 | GenBank |
| PX661558 | SriLanka | 2025-06-28 | GenBank |
| PX661559 | SriLanka | 2025-07-07 | GenBank |
| PX661560 | SriLanka | 2025-05-29 | GenBank |
| PX661561 | SriLanka | 2025-06-03 | GenBank |
| PX661562 | SriLanka | 2025-05-29 | GenBank |
| PX661563 | SriLanka | 2025-06-25 | GenBank |
| PX661564 | SriLanka | 2025-07-11 | GenBank |
| PX661565 | SriLanka | 2025-06-03 | GenBank |
| PX661566 | SriLanka | 2025-07-13 | GenBank |
| PX661567 | SriLanka | 2025-07-30 | GenBank |
| PX661568 | SriLanka | 2025-08-04 | GenBank |
| PX661569 | SriLanka | 2025-07-30 | GenBank |
| PX661570 | SriLanka | 2025-07-11 | GenBank |
| PX661571 | SriLanka | 2025-07-30 | GenBank |
| PX661572 | SriLanka | 2025-07-16 | GenBank |
| PX661573 | SriLanka | 2025-07-14 | GenBank |
| PX661574 | SriLanka | 2025-07-13 | GenBank |
| PX661575 | SriLanka | 2025-06-03 | GenBank |

**Supplementary Table 3. Public Genomes used for Mutational analysis (n = 59)**

| ID | Country | Collection_Date | Database_Type |
| --- | --- | --- | --- |
| PX661557 | SriLanka | 2025-06-08 | GenBank |
| PX661558 | SriLanka | 2025-06-28 | GenBank |
| PX661559 | SriLanka | 2025-07-07 | GenBank |
| PX661560 | SriLanka | 2025-05-29 | GenBank |
| PX661561 | SriLanka | 2025-06-03 | GenBank |
| PX661562 | SriLanka | 2025-05-29 | GenBank |
| PX661563 | SriLanka | 2025-06-25 | GenBank |
| PX661564 | SriLanka | 2025-07-11 | GenBank |
| PX661565 | SriLanka | 2025-06-03 | GenBank |
| PX661566 | SriLanka | 2025-07-13 | GenBank |
| PX661567 | SriLanka | 2025-07-30 | GenBank |
| PX661568 | SriLanka | 2025-08-04 | GenBank |
| PX661569 | SriLanka | 2025-07-30 | GenBank |
| PX661570 | SriLanka | 2025-07-11 | GenBank |
| PX661571 | SriLanka | 2025-07-30 | GenBank |
| PX661572 | SriLanka | 2025-07-16 | GenBank |
| PX661573 | SriLanka | 2025-07-14 | GenBank |
| PX661574 | SriLanka | 2025-07-13 | GenBank |
| PX661575 | SriLanka | 2025-06-03 | GenBank |
| GU013530.2 | SriLanka | 2008-04-01 | GenBank |
| FJ513645.1 | SriLanka | 2008-04-01 | GenBank |
| FJ513654.1 | SriLanka | 2008-04-01 | GenBank |
| FJ513657.1 | SriLanka | 2008-04-01 | GenBank |
| FJ513673.1 | SriLanka | 2008-04-01 | GenBank |
| FJ513675.1 | SriLanka | 2008-04-01 | GenBank |
| FJ513679.1 | SriLanka | 2008-04-01 | GenBank |
| FJ445426.2 | SriLanka | 2008-04-01 | GenBank |
| GU013528.2 | SriLanka | 2008-03-01 | GenBank |
| GU013529.2 | SriLanka | 2008-03-01 | GenBank |
| FJ513628.1 | SriLanka | 2008-03-01 | GenBank |
| FJ513629.1 | SriLanka | 2008-03-01 | GenBank |
| FJ513632.1 | SriLanka | 2008-03-01 | GenBank |
| FJ513635.1 | SriLanka | 2008-03-01 | GenBank |
| FJ513637.1 | SriLanka | 2008-03-01 | GenBank |
| FJ445427.2 | SriLanka | 2007-07-01 | GenBank |
| FJ445428.2 | SriLanka | 2007-05-01 | GenBank |
| HM045799.1 | SriLanka | 2007-01-01 | GenBank |
| HM045801.1 | SriLanka | 2007-01-01 | GenBank |
| MK086029.1 | SriLanka | 2006-11-14 | GenBank |
| GU189061.1 | SriLanka | 2006-01-01 | GenBank |
| EPI_ISL_19780906 | Brazil | 2025-02-11 | GISAID |
| EPI_ISL_19878101 | Brazil | 2025-01-30 | GISAID |
| EPI_ISL_20088612 | Mauritius | 2025-03-30 | GISAID |
| EPI_ISL_20090653 | Mauritius | 2025-04-16 | GISAID |
| EPI_ISL_20051872 | Brazil | 2025-04-22 | GISAID |
| EPI_ISL_20153716 | Brazil | 2025-03-07 | GISAID |
| PX216394.1 | China | 2025-07-31 | GenBank |
| PX216393.1 | China | 2025-07-27 | GenBank |
| PX216391.1 | China | 2025-07-24 | GenBank |
| EPI_ISL_19902404 | Mayotte | 2025-03-22 | GISAID |
| EPI_ISL_19902459 | Mayotte | 2025-04-12 | GISAID |
| PV685523.1 | Mayotte | 2025-04-11 | GenBank |
| PV685525.1 | Mayotte | 2025-03-29 | GenBank |
| EPI_ISL_19902278 | Mayotte | 2025-03-20 | GISAID |
| PV685695.1 | Reunion | 2025-04-02 | GenBank |
| PV685662.1 | Reunion | 2025-03-20 | GenBank |
| PV685675.1 | Reunion | 2025-03-13 | GenBank |
| PV685656.1 | Reunion | 2025-02-25 | GenBank |
| PV685667.1 | Reunion | 2025-02-10 | GenBank |

**Supplementary Table 4. GISAID acknowledgment table for sequences used in this study**

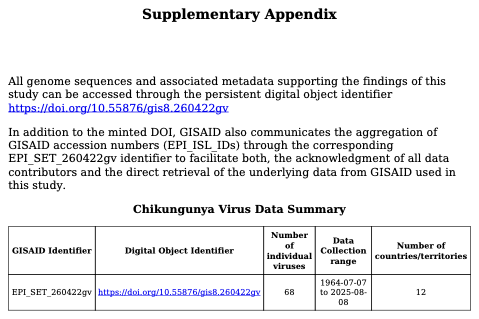

| ID: | **EPI_SET_260422gv** |
| --- | --- |
| DOI: | [**https://doi.org/10.55876/gis8.260422gv**](https://doi.org/10.55876/gis8.260422gv) |

**Supplementary Table 5. Public CHIKV genomes included in Time-scaled phylogeny and phylogeography of CHIKV IOL clade genomes. (n = 599)**

| ID | Location | Collection_Date | Included-in-171 seqs? |
| --- | --- | --- | --- |
| PX661557 | Sri_Lanka | 2025-06-08 | Yes |
| PX661558 | Sri_Lanka | 2025-06-28 | Yes |
| PX661559 | Sri_Lanka | 2025-07-07 | Yes |
| PX661560 | Sri_Lanka | 2025-05-29 | Yes |
| PX661561 | Sri_Lanka | 2025-06-03 | Yes |
| PX661562 | Sri_Lanka | 2025-05-29 | Yes |
| PX661563 | Sri_Lanka | 2025-06-25 | Yes |
| PX661564 | Sri_Lanka | 2025-07-11 | Yes |
| PX661565 | Sri_Lanka | 2025-06-03 | Yes |
| PX661575 | Sri_Lanka | 2025-06-03 | Yes |
| PX661566 | Sri_Lanka | 2025-07-13 | Yes |
| PX661567 | Sri_Lanka | 2025-07-30 | Yes |
| PX661568 | Sri_Lanka | 2025-08-04 | Yes |
| PX661569 | Sri_Lanka | 2025-07-30 | Yes |
| PX661570 | Sri_Lanka | 2025-07-11 | Yes |
| PX661571 | Sri_Lanka | 2025-07-30 | Yes |
| PX661572 | Sri_Lanka | 2025-07-16 | Yes |
| PX661573 | Sri_Lanka | 2025-07-14 | Yes |
| PX661574 | Sri_Lanka | 2025-07-13 | Yes |
| OQ918703.2 | South_Korea | 2022-12-28 | Yes |
| MH124570.2 | India | 2010 |  |
| PP349434.1 | India | 2006 | Yes |
| OR412350.1 | Malaysia | 2017-09-06 |  |
| OR412349.1 | Malaysia | 2017-07-21 |  |
| OR412348.1 | Malaysia | 2021-11-17 | Yes |
| OR412347.1 | Malaysia | 2021-12-24 |  |
| MW588419.1 | Malaysia | 2017-06-01 | Yes |
| MW588418.1 | Malaysia | 2017-06-13 |  |
| OR470611.1 | India | 2021-02-06 | Yes |
| OR470610.1 | India | 2021-02-06 |  |
| PP896920.1 | India | 2018 | Yes |
| PP896919.1 | India | 2018 |  |
| PP896918.1 | India | 2024 |  |
| PP896917.1 | India | 2024 |  |
| PP896916.1 | India | 2024 |  |
| PP896915.1 | India | 2024 |  |
| PP896914.1 | India | 2024 |  |
| PP896913.1 | India | 2024 |  |
| PP896912.1 | India | 2024 |  |
| PP896911.1 | India | 2024 |  |
| PP896910.1 | India | 2024 |  |
| PP896909.1 | India | 2024 |  |
| PP896908.1 | India | 2024 |  |
| PP896907.1 | India | 2024 |  |
| PP896906.1 | India | 2024 |  |
| PP896905.1 | India | 2024 |  |
| PP896904.1 | India | 2024 |  |
| PP896903.1 | India | 2024 |  |
| PP896902.1 | India | 2024 |  |
| PP896901.1 | India | 2024 |  |
| PP896900.1 | India | 2024 |  |
| PP896899.1 | India | 2024 |  |
| PP896898.1 | India | 2024 | Yes |
| PP896897.1 | India | 2024 |  |
| PP896896.1 | India | 2024 |  |
| PP896895.1 | India | 2024 |  |
| PP896894.1 | India | 2024 |  |
| PP626398.1 | Thailand | 2018 | Yes |
| PP626396.1 | Thailand | 2018 |  |
| PP599025.1 | Thailand | 2023-05-11 | Yes |
| OP168370.1 | Myanmar_Burma | 2019-07 | Yes |
| OP168369.1 | Myanmar_Burma | 2019-07 |  |
| OP168368.1 | Myanmar_Burma | 2019-07 |  |
| OP168367.1 | Myanmar_Burma | 2019-07 |  |
| OP168365.1 | Myanmar_Burma | 2019-07 |  |
| OP168363.1 | Myanmar_Burma | 2019-07 |  |
| OP168361.1 | Myanmar_Burma | 2019-07 |  |
| OP168360.1 | Myanmar_Burma | 2019-07 |  |
| OP168359.1 | Myanmar_Burma | 2019-07 |  |
| OP168356.1 | Myanmar_Burma | 2019-07 |  |
| OP168355.1 | Myanmar_Burma | 2019-07 |  |
| PP501554.1 | China | 2019 | Yes |
| PP501553.1 | China | 2019 |  |
| PP501552.1 | China | 2019 |  |
| OQ605454.1 | India | 2021 |  |
| OQ605453.1 | India | 2021 |  |
| OQ605452.1 | India | 2021 |  |
| OQ605451.1 | India | 2020 |  |
| OQ605450.1 | India | 2020 |  |
| OQ605449.1 | India | 2020 |  |
| OQ605448.1 | India | 2020 |  |
| OQ605447.1 | India | 2020 |  |
| OQ605446.1 | India | 2020 |  |
| OQ605445.1 | India | 2020 |  |
| OQ605442.1 | India | 2020 |  |
| OQ605441.1 | India | 2020 |  |
| OQ605440.1 | India | 2020 |  |
| OQ605439.1 | India | 2020 |  |
| OQ605438.1 | India | 2020 | Yes |
| OQ605437.1 | India | 2020 |  |
| OQ605436.1 | India | 2020 |  |
| OQ605435.1 | India | 2020 |  |
| OQ605434.1 | India | 2020 |  |
| PP193842.1 | Thailand | 2019-03-26 |  |
| PP193841.1 | Thailand | 2019-03-20 |  |
| PP193840.1 | Thailand | 2019-03-12 |  |
| PP193839.1 | Thailand | 2019-03-12 |  |
| PP193838.1 | Thailand | 2019-03-11 |  |
| PP193837.1 | Thailand | 2019-01-03 |  |
| PP193836.1 | Thailand | 2019-03-11 | Yes |
| PP193835.1 | Thailand | 2019-03-19 |  |
| PP193834.1 | Thailand | 2019-02-22 |  |
| PP193833.1 | Thailand | 2018-12-08 |  |
| PP193832.1 | Thailand | 2018-11-12 |  |
| PP236110.1 | Malaysia | 2021 | Yes |
| PP236109.1 | Malaysia | 2021 |  |
| PP236108.1 | Malaysia | 2021 |  |
| PP236107.1 | Malaysia | 2021 |  |
| PP236106.1 | Malaysia | 2021 |  |
| PP236105.1 | Malaysia | 2021 |  |
| OR715104.1 | India | 2023-10-17 | Yes |
| OR626603.1 | Saudi_Arabia | 2021 | Yes |
| OR037305.2 | China | 2017 |  |
| OR037309.1 | China | 2019 |  |
| OR037308.1 | China | 2019 | Yes |
| OR037307.1 | China | 2019 |  |
| OQ750694.1 | India | 2010 | Yes |
| OQ750692.1 | India | 2010 |  |
| OQ750691.1 | India | 2010 |  |
| OQ750690.1 | India | 2010 |  |
| OQ750689.1 | India | 2010 |  |
| OQ750688.1 | India | 2010 |  |
| ON887081.1 | India | 2022-05-28 | Yes |
| ON887080.1 | India | 2022-05-28 |  |
| ON887079.1 | India | 2022-05-28 |  |
| ON838557.1 | India | 2022-05-28 | Yes |
| ON838556.1 | India | 2022-05-28 |  |
| ON262796.1 | Thailand | 2019-11-14 |  |
| ON262795.1 | Thailand | 2019-11-07 |  |
| ON262794.1 | Thailand | 2019-10-17 |  |
| ON262793.1 | Thailand | 2019-10-15 |  |
| ON262792.1 | Thailand | 2012 | Yes |
| ON262791.1 | Thailand | 2012 |  |
| OL589270.1 | Cambodia | 2020-07-01 | Yes |
| MW557661.1 | Malaysia | 2020-09-03 | Yes |
| MW349426.1 | Thailand | 2020-10-01 | Yes |
| MW248364.1 | China | 2019-10-08 | Yes |
| MW248363.1 | China | 2019-10-10 |  |
| LC664166.1 | Thailand | 2020-09-25 | Yes |
| LC664165.1 | Thailand | 2020-09-13 |  |
| LC664164.1 | Thailand | 2020-09-08 |  |
| LC664163.1 | Thailand | 2020-08-28 |  |
| LC664162.1 | Thailand | 2020-08-26 |  |
| LC664161.1 | Thailand | 2020-08-20 |  |
| LC664160.1 | Thailand | 2020-08-19 |  |
| LC664159.1 | Thailand | 2020-08-04 |  |
| LC664158.1 | Thailand | 2020-07-21 |  |
| LC664157.1 | Thailand | 2020-06-29 |  |
| LC664156.1 | Maldives | 2019-08-02 |  |
| LC664155.1 | Maldives | 2019-06-18 |  |
| LC664154.1 | Maldives | 2019-05 |  |
| LC664153.1 | Maldives | 2019-05 |  |
| LC664152.1 | Maldives | 2019-05-19 |  |
| LC664151.1 | Maldives | 2019-04-14 |  |
| LC664150.1 | Maldives | 2019-04-10 |  |
| LC664149.1 | Maldives | 2019-04 |  |
| LC664148.1 | Maldives | 2019-04-04 |  |
| LC664147.1 | Maldives | 2019-04-07 |  |
| LC664146.1 | Maldives | 2019-04-03 |  |
| LC664145.1 | Maldives | 2019-04-01 |  |
| LC664144.1 | Maldives | 2019-04-24 |  |
| LC664143.1 | Maldives | 2019-03-30 |  |
| LC664142.1 | Maldives | 2019-03-31 |  |
| LC664141.1 | Maldives | 2019-03-30 |  |
| OL999095.1 | Cambodia | 2021-05-24 | Yes |
| OL999094.1 | Cambodia | 2021-03-05 |  |
| OL999093.1 | Cambodia | 2021-02-24 |  |
| OL999092.1 | Cambodia | 2021-02-12 |  |
| OL999091.1 | Cambodia | 2020-12-25 |  |
| OL979156.1 | Cambodia | 2020-11-16 | Yes |
| OL979155.1 | Cambodia | 2020-11-18 |  |
| OL979154.1 | Cambodia | 2020-12-07 |  |
| OL979153.1 | Cambodia | 2020-12-24 |  |
| OL893107.1 | Cambodia | 2020-10-14 |  |
| OL893106.1 | Cambodia | 2020-10-09 | Yes |
| OL893105.1 | Cambodia | 2020-10-06 |  |
| OL849991.1 | Cambodia | 2020-09-28 |  |
| OL849990.1 | Cambodia | 2020-09-22 | Yes |
| OL741630.1 | Cambodia | 2020-07-23 | Yes |
| OL705486.1 | Cambodia | 2020-07-15 | Yes |
| MZ443816.1 | Thailand | 2018-08 | Yes |
| MZ443815.1 | Thailand | 2018-08 |  |
| MZ443814.1 | Thailand | 2018-08 |  |
| MZ443813.1 | Thailand | 2018-08 |  |
| MW291576.1 | China | 2019-10-04 | Yes |
| MT668625.1 | China | 2019-11-04 | Yes |
| MH636708.1 | Singapore | 2010-01 | Yes |
| MW574902.1 | India | 2016 | Yes |
| MW110477.1 | China | 2019-10-12 | Yes |
| MW110476.1 | China | 2019-10-12 |  |
| MW110475.1 | China | 2019-10-08 |  |
| MW110474.1 | China | 2019-10-06 |  |
| MW110473.1 | China | 2019-10-06 |  |
| MW110472.1 | China | 2019-09-28 |  |
| MT640256.1 | Thailand | 2020-02-05 |  |
| MT640255.1 | Thailand | 2020-01-17 | Yes |
| MT495608.1 | Thailand | 2019-11-06 |  |
| MT495607.1 | Thailand | 2019-08-28 |  |
| MT495606.1 | Thailand | 2019-08-12 | Yes |
| MT495605.1 | Thailand | 2019-04-14 |  |
| MW321606.1 | India | 2016 | Yes |
| MW042255.1 | India | 2019-06 | Yes |
| MW042254.1 | India | 2014-10 |  |
| MT123010.1 | China | 2017-09-01 | Yes |
| MT123009.1 | China | 2017-09-01 |  |
| MT123008.1 | China | 2017-09-01 |  |
| MT023791.1 | Djibouti | 2019-12-15 | Yes |
| LC580270.1 | Thailand | 2010 | Yes |
| LC580269.1 | Thailand | 2019-10 |  |
| LC580268.1 | Thailand | 2019-10 |  |
| LC580267.1 | Thailand | 2019-10 |  |
| LC580266.1 | Thailand | 2019-10 |  |
| LC580265.1 | Thailand | 2019-10 |  |
| LC580264.1 | Thailand | 2019-10 |  |
| LC580263.1 | Thailand | 2019-10 |  |
| LC580262.1 | Thailand | 2019-10 |  |
| LC580261.1 | Thailand | 2019-10 |  |
| LC580260.1 | Thailand | 2019-10 |  |
| LC580259.1 | Thailand | 2019-10 |  |
| LC580258.1 | Thailand | 2019-10 |  |
| LC580257.1 | Thailand | 2019-10 |  |
| LC580256.1 | Thailand | 2019-10 |  |
| LC580255.1 | Bangladesh | 2017-12 |  |
| LC580254.1 | Bangladesh | 2017-11 |  |
| LC580253.1 | Bangladesh | 2017-11 |  |
| LC580252.1 | Bangladesh | 2017-10 |  |
| LC580251.1 | Bangladesh | 2017-10 |  |
| LC580250.1 | Bangladesh | 2017-10 |  |
| LC580249.1 | Bangladesh | 2017-10 |  |
| LC580248.1 | Bangladesh | 2017-10 |  |
| LC580247.1 | Bangladesh | 2017-09 |  |
| LC580246.1 | Bangladesh | 2017-09 |  |
| LC580245.1 | Bangladesh | 2017-09 |  |
| LC580244.1 | Bangladesh | 2017-09 |  |
| LC580243.1 | Bangladesh | 2017-09 |  |
| LC580242.1 | Bangladesh | 2017-09 |  |
| LC580241.1 | Bangladesh | 2017-09 |  |
| LC580240.1 | Bangladesh | 2017-08 |  |
| LC580239.1 | Bangladesh | 2017-07 |  |
| LC580238.1 | Bangladesh | 2017-07 |  |
| LC580237.1 | Bangladesh | 2017-07 |  |
| LC580236.1 | Bangladesh | 2017-07 |  |
| MT380161.1 | Kenya | 2017-12 | Yes |
| MT380160.1 | Kenya | 2018-04 |  |
| MT380159.1 | Kenya | 2018-05 |  |
| MT380155.1 | Kenya | 2018-04 |  |
| MT380154.1 | Kenya | 2018-04 |  |
| MT380153.1 | Kenya | 2018-01 |  |
| MT380152.1 | Kenya | 2018-01 |  |
| MT380151.1 | Kenya | 2018-01 |  |
| MT380150.1 | Kenya | 2018-01 |  |
| MT380149.1 | Kenya | 2018-01 |  |
| MT380148.1 | Kenya | 2018-01 |  |
| MT380147.1 | Kenya | 2018-02 |  |
| MT380146.1 | Kenya | 2018-02 |  |
| MH647218.1 | Singapore | 2013-03 | Yes |
| MH647217.1 | Singapore | 2013-02 |  |
| MH647216.1 | Singapore | 2010-01 |  |
| MH647215.1 | Singapore | 2010-02 |  |
| MH647214.1 | Singapore | 2010-01 |  |
| MH647213.1 | Singapore | 2010-01 |  |
| MH647212.1 | Singapore | 2009-11 |  |
| MH647211.1 | Singapore | 2009-12 |  |
| MH647210.1 | Singapore | 2009-12 |  |
| MH647209.1 | Singapore | 2013-06 |  |
| MH647208.1 | Singapore | 2017-06 |  |
| MH647207.1 | Singapore | 2015-10 |  |
| MH647206.1 | Singapore | 2014-06 |  |
| MH647205.1 | Singapore | 2014-04 |  |
| MH647204.1 | Singapore | 2014-02 |  |
| MH647203.1 | Singapore | 2014-02 |  |
| MH647202.1 | Singapore | 2014-01 |  |
| MH647201.1 | Singapore | 2013-12 |  |
| MH647200.1 | Singapore | 2013-12 |  |
| MH647199.1 | Singapore | 2013-12 | Yes |
| MH647198.1 | Singapore | 2013-10 |  |
| MH647197.1 | Singapore | 2013-10 |  |
| MH647196.1 | Singapore | 2013-08 |  |
| MH647195.1 | Singapore | 2013-07 |  |
| MH647194.1 | Singapore | 2013-06 |  |
| MH647192.1 | Singapore | 2013-05 |  |
| MH647191.1 | Singapore | 2013-05 |  |
| MH647190.1 | Singapore | 2013-05 |  |
| MH647189.1 | Singapore | 2013-04 |  |
| MH647188.1 | Singapore | 2013-04 |  |
| MH647187.1 | Singapore | 2013-02 |  |
| MH647186.1 | Singapore | 2012-11 |  |
| MH647184.1 | Singapore | 2012-10 |  |
| MH647182.1 | Singapore | 2012-01 |  |
| MH647181.1 | Singapore | 2011-11 |  |
| MH647180.1 | Singapore | 2011-10 |  |
| MN756625.1 | China | 2019-08-08 | Yes |
| MT526796.1 | Kenya | 2018-01-15 | Yes |
| MH349097.1 | China | 2017-04-30 | Yes |
| MK040571.1 | Thailand | 2018-08-21 | Yes |
| MK040570.1 | Thailand | 2018-08-24 |  |
| MK040569.1 | Thailand | 2018-08-25 |  |
| MN974224.1 | Thailand | 2019-09-06 |  |
| MN974213.1 | Thailand | 2019-10-07 |  |
| MN974212.1 | Thailand | 2019-07-08 |  |
| MN974211.1 | Thailand | 2018-12-12 |  |
| MN974210.1 | Thailand | 2019-08-26 | Yes |
| MN974209.1 | Thailand | 2019-10-07 |  |
| MN974208.1 | Thailand | 2019-02-12 |  |
| MN974207.1 | Thailand | 2019-03-12 |  |
| MN974206.1 | Thailand | 2019-08-30 |  |
| MN974205.1 | Thailand | 2019-07-24 |  |
| MN974204.1 | Thailand | 2019-08-15 |  |
| MN974203.1 | Thailand | 2019-09-11 |  |
| MN974223.1 | Thailand | 2018-07-21 |  |
| MN974222.1 | Thailand | 2018-07-21 |  |
| MN974221.1 | Thailand | 2018-07-19 |  |
| MN974220.1 | Thailand | 2018-07-19 |  |
| MN974219.1 | Thailand | 2018-07-13 |  |
| MN974218.1 | Thailand | 2018-07-02 |  |
| MN974217.1 | Thailand | 2018-07-02 |  |
| MN974216.1 | Thailand | 2018-06-30 |  |
| MN974215.1 | Thailand | 2018-06-23 |  |
| MN974214.1 | Thailand | 2018-06-20 |  |
| MN075150.1 | Finland | 2019-02 | Yes |
| MN075149.1 | Finland | 2019-02 |  |
| MK370033.1 | India | 2016 |  |
| MK370032.1 | India | 2016 |  |
| MK370031.1 | India | 2015 |  |
| MK370030.1 | India | 2015 | Yes |
| MN402892.1 | China | 2019-08-02 | Yes |
| MN402891.1 | China | 2019-07-30 |  |
| MN402890.1 | China | 2019-07-26 |  |
| MN402889.1 | China | 2019-07-25 |  |
| MN402888.1 | Myanmar_Burma | 2019-08-01 |  |
| MN402887.1 | Myanmar_Burma | 2019-08-01 |  |
| MN402886.1 | China | 2019-07-02 |  |
| MN402885.1 | China | 2019-05-08 |  |
| MN402884.1 | China | 2019-06-28 |  |
| MN402883.1 | China | 2019-05-07 |  |
| MK286899.1 | India | 2017 | Yes |
| MK286898.1 | India | 2018 |  |
| MK286897.1 | India | 2018 |  |
| MK286896.1 | India | 2017 |  |
| MK286895.1 | India | 2014 |  |
| MK286894.1 | India | 2017 |  |
| MK286893.1 | India | 2017 |  |
| MH124582.1 | India | 2016 | Yes |
| MH124581.1 | India | 2016 |  |
| MH124580.1 | India | 2016 |  |
| MH124579.1 | India | 2010 |  |
| MH124578.1 | India | 2010 |  |
| MH124575.1 | India | 2010 |  |
| MH124573.1 | India | 2010 |  |
| MH124572.1 | India | 2010 |  |
| MH124571.1 | India | 2010 |  |
| MK468621.1 | Bangladesh | 2017-07-18 |  |
| MK468620.1 | Bangladesh | 2017-07-11 |  |
| MK468618.1 | Bangladesh | 2017-06-29 |  |
| MK468617.1 | Bangladesh | 2017-06-21 |  |
| MK468615.1 | Bangladesh | 2017-07-10 |  |
| MK468613.1 | Bangladesh | 2017-06-24 |  |
| MK468612.1 | Bangladesh | 2017-06-17 |  |
| MK468611.1 | Bangladesh | 2017-05-31 |  |
| MK468610.1 | Bangladesh | 2017-11-22 |  |
| MK468609.1 | Bangladesh | 2017-06-19 |  |
| MK468608.1 | Bangladesh | 2017-06-17 | Yes |
| MN630017.1 | Australia | 2019 | Yes |
| MK468801.1 | Thailand | 2018-06-27 | Yes |
| MK551553.1 | India | 2016-10-03 | Yes |
| MK551552.1 | India | 2016-09-30 |  |
| MK518340.1 | India | 2016-09-28 | Yes |
| MK028838.1 | Sri_Lanka | 2006 | Yes |
| MK028836.1 | Comoros | 2005 | Yes |
| MK848202.1 | Thailand | 2018-11-21 | Yes |
| MK086029.1 | Sri_Lanka | 2006-11-14 | Yes |
| MK473631.1 | India | 2016-09-11 |  |
| MK473630.1 | India | 2016 |  |
| MK473629.1 | India | 2016-10-18 | Yes |
| MK473628.1 | India | 2016-10-13 |  |
| MK473627.1 | India | 2016-10-23 |  |
| MK473626.1 | India | 2016-10-06 |  |
| MK473625.1 | India | 2016-08-26 |  |
| MK473624.1 | India | 2016-10-23 |  |
| MK473623.1 | India | 2016-09-20 |  |
| MK473622.1 | India | 2016-09-13 |  |
| MK473621.1 | India | 2016-09-02 |  |
| MK120202.1 | Italy | 2007-08-25 | Yes |
| MK120201.1 | Italy | 2007-08-20 |  |
| MK120200.1 | Italy | 2017-08-09 |  |
| MK120199.1 | Italy | 2017-08-02 | Yes |
| MK120198.1 | Italy | 2017-08-02 |  |
| MK120197.1 | Italy | 2017-08-02 |  |
| MK120196.1 | Italy | 2017-09-30 |  |
| MK120195.1 | Italy | 2017-09-27 |  |
| MK120194.1 | Italy | 2017-09-27 |  |
| MH754507.1 | Italy | 2017-09-08 | Yes |
| MH423810.1 | Kenya | 2016-05-31 | Yes |
| MH423806.1 | Kenya | 2016-06-01 |  |
| MH423805.1 | Kenya | 2016-06-01 |  |
| MH423804.1 | Kenya | 2016-06-01 |  |
| MH423803.1 | Kenya | 2016-05-31 |  |
| MH423802.1 | Kenya | 2016-05-31 |  |
| MH423801.1 | Kenya | 2016-05-31 |  |
| MH423800.1 | Kenya | 2016-05-31 |  |
| MH423799.1 | Kenya | 2016-05-31 | Yes |
| MH423798.1 | Kenya | 2016-05-08 |  |
| MH423797.1 | Kenya | 2016-05-07 |  |
| MH400249.1 | China | 2017-08-23 | Yes |
| MH507158.1 | Italy | 2017 | Yes |
| MH229986.1 | Mauritius | 2006 | Yes |
| MG912993.1 | China | 2017-08-30 | Yes |
| MG137428.1 | India | 2016-06-29 | Yes |
| KT324228.1 | Malaysia | 2009-02 | Yes |
| KT324227.1 | Malaysia | 2009-03 |  |
| KT324226.1 | Malaysia | 2009-01 |  |
| KT324225.1 | Malaysia | 2009-01 |  |
| KT324224.1 | Malaysia | 2008-12 |  |
| MF740874.1 | Pakistan | 2017 | Yes |
| MG664850.1 | China | 2010 | Yes |
| KX619423.1 | India | 2014-07-23 |  |
| KX619422.1 | India | 2014-07-08 |  |
| MG925665.1 | China | 2017-12-15 | Yes |
| MF773569.1 | Papua_New_Guinea | 2013 | Yes |
| MF773568.1 | Malaysia | 2008 |  |
| MF773566.1 | Bangladesh | 2017 |  |
| LC259093.1 | Malaysia | 2009-01-06 | Yes |
| LC331252.1 | Japan | 2016 | Yes |
| MG049915.1 | Italy | 2017 | Yes |
| MF076577.1 | Laos | 2012-08-31 | Yes |
| MF076576.1 | Laos | 2012-08-31 |  |
| MF076575.1 | Laos | 2012-08-16 |  |
| MF076574.1 | Laos | 2012-08-16 |  |
| MF076573.1 | Laos | 2012-08-07 |  |
| MF076572.1 | Laos | 2013-03-12 |  |
| MF076571.1 | Laos | 2013-03-28 |  |
| MF076570.1 | Laos | 2013-03-28 |  |
| MF076569.1 | Laos | 2013-03-28 |  |
| MF076568.1 | Laos | 2013-03-28 |  |
| MF503628.1 | Hong_Kong_SAR_China | 2016-08-26 | Yes |
| MF499120.1 | Hong_Kong_SAR_China | 2016-09-15 | Yes |
| MF774619.1 | Pakistan | 2016 | Yes |
| MF774618.1 | Pakistan | 2016 |  |
| MF774617.1 | Pakistan | 2016 |  |
| MF774616.1 | Pakistan | 2016 |  |
| MF774615.1 | Pakistan | 2016 |  |
| MF774614.1 | Pakistan | 2016 |  |
| MF774613.1 | Pakistan | 2016 |  |
| KX619425.1 | India | 2015-09-11 | Yes |
| KX619424.1 | India | 2015-09-22 |  |
| KY575571.1 | United_States | 2006 | Yes |
| KY575570.1 | United_States | 2008 |  |
| KY575568.1 | United_States | 2006 |  |
| KY575567.1 | United_States | 2006 |  |
| KU365371.1 | Bangladesh | 2011-11 |  |
| KU365370.1 | Bangladesh | 2011-11 | Yes |
| KY751908.1 | Australia | 2016 | Yes |
| KX009171.1 | Thailand | 2013 | Yes |
| KX009170.1 | Thailand | 2013 |  |
| KX009169.1 | Thailand | 2013 |  |
| KX009168.1 | Thailand | 2013 |  |
| KX009167.1 | Thailand | 2013 |  |
| KY057363.1 | India | 2016-08-28 | Yes |
| KX262997.1 | Malaysia | 2009 | Yes |
| KX262993.1 | Italy | 2007 |  |
| KX262989.1 | Italy | 2007 |  |
| FJ445511.2 | Singapore | 2008-01 |  |
| FJ445510.2 | Singapore | 2008-01 |  |
| FJ445502.2 | Singapore | 2008-08 | Yes |
| FJ445484.2 | Singapore | 2008-05 |  |
| FJ445463.2 | Singapore | 2008-07 |  |
| FJ445445.2 | Singapore | 2008-08 |  |
| FJ445443.2 | Singapore | 2008-08 |  |
| FJ445433.2 | Singapore | 2008-08 |  |
| FJ445432.2 | Singapore | 2008-07 |  |
| FJ445431.2 | Singapore | 2008-07 |  |
| FJ445430.2 | Singapore | 2008-07 |  |
| FJ445428.2 | Sri_Lanka | 2007-05 | Yes |
| FJ445427.2 | Sri_Lanka | 2007-07 | Yes |
| FJ445426.2 | Sri_Lanka | 2008-04 | Yes |
| FJ513679.1 | Sri_Lanka | 2008-04 | Yes |
| FJ513675.1 | Sri_Lanka | 2008-04 | Yes |
| FJ513673.1 | Sri_Lanka | 2008-04 | Yes |
| FJ513657.1 | Sri_Lanka | 2008-04 | Yes |
| FJ513654.1 | Sri_Lanka | 2008-04 | Yes |
| FJ513645.1 | Sri_Lanka | 2008-04 | Yes |
| FJ513637.1 | Sri_Lanka | 2008-03 | Yes |
| FJ513635.1 | Sri_Lanka | 2008-03 | Yes |
| FJ513632.1 | Sri_Lanka | 2008-03 | Yes |
| FJ513629.1 | Sri_Lanka | 2008-03 | Yes |
| FJ513628.1 | Sri_Lanka | 2008-03 | Yes |
| FR687348.1 | Malaysia | 2009-04-03 |  |
| FR687347.1 | Malaysia | 2009-02-12 |  |
| FR687346.1 | Malaysia | 2009-02-16 | Yes |
| FR687345.1 | Malaysia | 2009-02-05 |  |
| FR687344.1 | Malaysia | 2009-01-12 |  |
| FR687343.1 | Malaysia | 2008-12-12 |  |
| FR687342.1 | Malaysia | 2008-11-10 |  |
| FR687341.1 | Malaysia | 2008-11-05 |  |
| FR687340.1 | Malaysia | 2008-08-21 |  |
| KP702297.1 | Comoros | 2005 | Yes |
| KF283987.1 | Comoros | 2005 | Yes |
| KT336782.1 | India | 2013-05-27 |  |
| KT336781.1 | India | 2013-03-06 |  |
| KT336780.1 | India | 2012-08-03 |  |
| KT336779.1 | India | 2012-08-03 |  |
| KT336778.1 | India | 2010-07-16 |  |
| KT336777.1 | India | 2009-04-08 | Yes |
| KC614648.1 | Yemen | 2011-01-25 | Yes |
| KP164869.1 | Thailand | 2009 | Yes |
| KF151175.1 | Myanmar_Burma | 2009-12-11 | Yes |
| KF151174.1 | Myanmar_Burma | 2009-07-13 |  |
| KP003811.1 | Italy | 2007 |  |
| KP003810.1 | Italy | 2007 |  |
| KP003809.1 | Mayotte | 2006 |  |
| KP003808.1 | Madagascar | 2006 | Yes |
| KP003807.1 | France | 2006 |  |
| KJ579187.1 | Thailand | 2013-10-14 | Yes |
| KJ579186.1 | Thailand | 2013-10-14 |  |
| KJ579185.1 | Thailand | 2013-10-14 |  |
| KJ579184.1 | Thailand | 2013-10-14 |  |
| KF590567.1 | Myanmar_Burma | 2010 | Yes |
| KF590566.1 | Myanmar_Burma | 2010 |  |
| KF590565.1 | Myanmar_Burma | 2010 |  |
| KF590564.1 | Myanmar_Burma | 2010 |  |
| KJ941050.1 | United_States | 2006 | Yes |
| KJ796852.1 | Thailand | 2009-02-12 |  |
| KJ796851.1 | Thailand | 2008-12-01 |  |
| KJ796850.1 | Thailand | 2009-09-16 |  |
| KJ796849.1 | Thailand | 2008-12-23 |  |
| KJ796848.1 | Thailand | 2008-11-06 |  |
| KJ796847.1 | Thailand | 2009-03-05 |  |
| KJ796846.1 | India | 2009-08-18 | Yes |
| KJ796845.1 | India | 2009-08-04 |  |
| KJ796844.1 | India | 2009-08-04 |  |
| JQ065892.1 | China | 2010-10 |  |
| JQ065891.1 | China | 2010-10 |  |
| JQ065890.1 | China | 2010-10 |  |
| JQ065889.1 | China | 2010-10 |  |
| JQ065888.1 | China | 2010-10 |  |
| JQ065887.1 | China | 2010-10 |  |
| JQ065886.1 | China | 2010-10 | Yes |
| JQ065885.1 | China | 2010-10 |  |
| KC862329.1 | Indonesia | 2010 | Yes |
| HQ846359.1 | China | 2010-10 | Yes |
| HQ846358.1 | China | 2010-10 |  |
| HQ846357.1 | China | 2010-10 |  |
| HQ846356.1 | China | 2010-10 |  |
| JN558836.1 | India | 2009 | Yes |
| JN558835.1 | India | 2008 |  |
| JN558834.1 | India | 2009 |  |
| FN295485.3 | Malaysia | 2008 | Yes |
| FN295487.2 | Malaysia | 2008 |  |
| JQ861260.1 | Cambodia | 2011-05-28 |  |
| JQ861259.1 | Cambodia | 2011-05-26 |  |
| JQ861258.1 | Cambodia | 2011-08-16 | Yes |
| JQ861257.1 | Cambodia | 2011-08-16 |  |
| JQ861256.1 | Cambodia | 2011-08-16 |  |
| JQ861255.1 | Cambodia | 2011-08-16 |  |
| JQ861254.1 | Cambodia | 2011-08-16 |  |
| JQ861253.1 | Cambodia | 2011-08-16 |  |
| FR717337.1 | Reunion | 2005-12-26 | Yes |
| FR717336.1 | Reunion | 2005-12-26 |  |
| JF274082.1 | India | 2006-09-27 | Yes |
| JX088705.1 | China | 2010 | Yes |
| GU189061.1 | Sri_Lanka | 2006 | Yes |
| HM045801.1 | Sri_Lanka | 2007 | Yes |
| HM045799.1 | Sri_Lanka | 2007 | Yes |
| HM045794.1 | United_States | 2006 | Yes |
| GU301781.1 | Thailand | 2009-07-27 |  |
| GU301780.1 | Thailand | 2008-10-21 |  |
| GU301779.1 | Thailand | 2009-09-04 | Yes |
| GU908223.1 | Thailand | 2009-08-14 | Yes |
| GQ905863.1 | Thailand | 2009-05-25 | Yes |
| GU199353.1 | China | 2008 |  |
| GU199352.1 | China | 2008 |  |
| GU199351.1 | China | 2008 | Yes |
| GU199350.1 | China | 2008 |  |
| GU013528.2 | Sri_Lanka | 2008-03 | Yes |
| GU013529.2 | Sri_Lanka | 2008-03 | Yes |
| GU013530.2 | Sri_Lanka | 2008-04 | Yes |
| FJ807899.1 | Malaysia | 2008 |  |
| FJ807898.1 | Bangladesh | 2008 | Yes |
| FJ807896.1 | Singapore | 2006 |  |
| GQ428215.1 | India | 2008-05-29 | Yes |
| GQ428214.1 | India | 2008-06-29 |  |
| GQ428213.1 | India | 2007-07-13 |  |
| GQ428212.1 | India | 2007-07-12 |  |
| GQ428211.1 | India | 2006-10-07 |  |
| GQ428210.1 | India | 2006-10-07 |  |
| FJ959103.1 | Mauritius | 2006 | Yes |
| FJ000069.1 | India | 2007-06 |  |
| FJ000068.1 | India | 2006-08 |  |
| FJ000067.1 | India | 2006-08 | Yes |
| FJ000066.1 | India | 2006-09 |  |
| FJ000065.1 | India | 2006-09 |  |
| FJ000064.1 | India | 2006-09 |  |
| FJ000063.1 | India | 2006-10 |  |
| FJ000062.1 | India | 2006-09 |  |
| EU244823.2 | Italy | 2007 | Yes |
| EU372006.1 | India | 2007-06-11 | Yes |
| EF210157.2 | India | 2006 | Yes |
| EU564335.1 | India | 2006-10-31 |  |
| EU564334.1 | Mauritius | 2006-02-14 | Yes |
